# Mitigation Policies and Emergency Care Management in Europe’s Ground Zero for COVID-19^*^

**DOI:** 10.1101/2020.05.19.20106575

**Authors:** Gabriele Ciminelli, Sílvia Garcia-Mandicó

## Abstract

This paper draws from daily death registry data on 4,000 Italian municipalities to investigate two crucial policies that can dramatically affect the toll of COVID-19: the shutdown of non-essential businesses and the management of the emergency care system. Our results, which are robust to controlling for a host of co-factors, offer strong evidence that the closure of service activities is very effective in reducing COVID-19 mortality – this was about 15% lower in municipalities with a 10 percentage points higher employment share in shut down services. Shutting down factories, instead, is much less effective, plausibly because factory workers engage in more limited physical interactions relative to those in the consumer-facing service sector. Concerning the management of the health care system, we find that mortality strongly increases with distance from the intensive care unit (ICU). Municipalities at 10 km from the closest ICU experienced up to 50% higher mortality. This effect – which is largest within the epicenter and in days of abnormally high volumes of calls to the emergency line – underscores the importance of improving pre-hospital emergency services and building ambulance capacity to ensure timely transportation of critical patients to the ICU.

## 1 Introduction

Containment and mitigation strategies, such as travel restrictions, people lockdowns, and business shutdowns, aim at reducing the pace and extent of COVID-19 infections (“flattening the curve”). These measures are thought to save lives by alleviating the burden on health care systems, thus allowing the recovery of patients (Ferguson et al., 2020). However, if mitigation policies are not effective quickly enough in flattening the curve, having a prepared and responsive health care system is the only way to preventing a runaway death toll. This paper focuses on two policies in particular – the shutdowns of non-essential business and the management of the emergency care system – and analyses what could be done better should a second wave materialize.

Our focus is on Italy, which offers a very good case study (Briscese et al., 2020). Being among the first countries to be struck by COVID-19, its government had to implement hastily arranged mitigation policies, including the shutdown of all non-essential business. As many countries either adopted Italy’s policies or are considering to do so, investigating their effectiveness is very relevant for policymakers around the world. At the same time, the high level of contagion observed in Italy brought its health care system close to collapse (Johnson, 2020). Quantifying how much better preparedness could have helped in reducing COVID-19 mortality thus offers valuable lessons for any country at risk of being struck by COVID-19.

Focusing on Italy also has another key advantage in our context – the availability of highly granular daily death registry data for thousands of municipalities. These data allow for a more precise estimation of the effect of COVID-19 on the mortality rate relative to official COVID-19 fatality data, which have serious undercounting issues, as we document below (see also Muellbauer and Aron, 2020). Besides death registry data, Italy also makes available several other useful municipality-level datasets. We have detailed health care data, as well as data on employment shares in sectors shutdown by the government and on a host of other co-factors, such as population density, commuting, digital employment, air pollution, and several demographic characteristics. This extraordinary wealth of data, which to our knowledge is unmatched in other countries, allows us to exploit exogenous variation in underlining municipality characteristics to credibly isolate the effect of government policies on mortality outcomes, while also controlling for many other possible determinants of COVID-19 mortality.

Our focus is on 4,000 municipalities in Italy’s north, accounting for about 50% of the overall population and about 85% of all official COVID-19 fatalities. We start the analysis by establishing some key stylized facts. The data indicate that the epidemic may have induced the death of almost 0.1% of the local population in just over a month (from February/21^st^ to March/31^st^ of 2020), and that its mortality is vastly undercounted in official statistics. A conservative estimate suggests that an additional 1.2 deaths went undetected for each officially recorded COVID-19 fatality. We then estimate the effect of COVID-19 on mortality. Our results highlight important heterogeneities, both across regions and over time. COVID-19 killed an average of two people per day for each 100,000 inhabitants. But within the outbreak epicenter its mortality effect peaked at twelve deaths per day for each 100,000 inhabitants.

In the second part of the paper, we analyze the effectiveness of business shutdowns in reducing mortality. Since about one quarter of COVID-19 infections occurs through the workplace (Lewandowski et al., 2020; OECD, 2020), workplace social distancing can potentially be a very effective mitigation measure. Yet, it is also an extremely costly policy, which governments may not want to apply across the board (del Rio-Chanona et al., 2020; Koren and Pető, 2020). In Italy, the government ordered the closure of all firms in specific non-essential sectors at the national level. We exploit variation in the share of employment in suspended sectors across municipalities to identify the effect of the government policy, following Rajan and Zingales (1998).

Our results, which are robust to controlling for many other co-factors, offer strong evidence that the closure of service activities was very effective in reducing COVID-19 mortality – this was about 15% lower in municipalities with a 10 percentage points higher share of employment in close down services. On the other hand, our results suggest that shutting down factories may have not been that effective. A plausible explanation for this difference is that factory workers engage in fairly limited physical interactions relative to workers in the consumer-facing service sector. Our results suggest that governments should not hesitate to close down services in the quest of halting COVID-19 mortality, but they should more carefully weight the less clear benefits of closing down factories against the undoubted costs given by the halt in production.

In the final part of the paper, we zoom in on Italy’s COVID-19 outbreak epicenter and assess whether the burden on its health care system contributed to the high COVID-19 mortality. In particular, we focus on pre-hospital emergency care (emergency line response and ambulance availability). The timely arrival of patients to intensive care units (ICU) is largely determined by the competence of pre-hospital emergency care. As COVID-19 spreads among the population, calls to the emergency line soar, and waiting times for emergency transportation swell, with ambulances often being unable to get in on time (Sorbi, 2020; New York Times, 2020). Such bottlenecks can raise cumulative mortality, undermining countries’ efforts to build ICU capacity. We explore whether a higher distance to the ICU may have resulted in higher mortality, at times of burdened emergency transportation.

Strikingly, we find that COVID-19 had a significant effect on mortality for people as young as 40 years old in municipalities far from an ICU. Among those above 55, municipalities with an ICU in town consistently experienced 30% to 50% lower mortality rates. These results – which are very robust and are not observed for areas outside the epicenter – suggest that, given resource and time constraints, medical staff may have had to prioritize serving more patients at the expense of reducing geographical coverage. This highlights the importance of increasing preparedness – both in Italy and abroad – to help reduce mortality, shall new outbreaks materialize. Governments should improve pre-hospital emergency services, build ambulance capacity, and, ideally, mobilize ICUs more evenly across the territory.

This paper contributes to a burgeoning literature quantifying the effectiveness of social distancing policies on COVID-19 mortality. A large part of this literature has focused on evaluating the effects of people lockdowns and travel restrictions on contagion, and ultimately, mortality (Adda, 2016; Becchetti et al., 2020; Chinazzi et al., 2020; Fang et al., 2020; Ferguson et al., 2020; Juranek and Zoutman, 2020; Pedersen and Meneghini, 2020). Our focus is instead on the effect of businesses shutdowns. To the best of our knowledge, we are the first to quantify the effectiveness of such policies. Our results relate to Lewandowski et al. (2020) and Muellbauer and Aron (2020). The former find that physical contact through occupational exposure can explain up to a quarter of the spread and mortality of COVID-19 across Europe. The latter find that most of COVID-19 deaths in England were among people employed in the consumer-facing service sector. Finally, we also relate to the literature that seeks to understand the causes behind Italy’s severe COVID-19 outbreak (see, for example, Gatto et al., 2020; Pluchino et al., 2020; Rinaldi and Paradisi, 2020). We contribute to this strand of work by uncovering the congestion of the health care system as an important reason behind its high death toll.

The rest of the paper is organized as follows: Section 2 describes the dataset, while in Section 3 we summarize four main stylized facts that emerge from the data. Section 4 briefly describes the methodology and presents the main effects of COVID-19 on mortality. In Section 5 we assess the effectiveness of business shutdowns in curbing COVID-19 mortality. Section 6 zooms in on the outbreak epicenter and explores the management of the emergency care system. Section 7 concludes.

## 2 Dataset

We source death registry data at the municipality-level from ISTAT (2020c), the Italian statistical agency. The data provide information on daily deaths by age and gender for the January/1^st^-March/31^st^ period, for the years 2015 to 2020.^1^ Our focus is on municipalities in the eight regions of Italy’s north – Emilia-Romagna, Friuli-Venezia Giulia, Liguria, Lombardia, Piemonte, Trentino-Alto Adige, Valle d’Aosta, and Veneto – which together account for about 50% and 85% of respectively Italy’s population and its official COVID-19 fatalities, as measured by Protezione Civile (2020). The sample covers 4,000 municipalities, almost all in the eight regions that we consider. Table A1 in Appendix A summarizes sample statistics.

To compute mortality rates, we source census data on population at the municipality-level, by age and gender, from ISTAT (2020*d*). The dataset is then complemented by variables capturing slow-moving municipality characteristics that we use to explore potential co-factors of COVID-19 mortality. These characteristics broadly fall in four categories: (i) socio-demographic characteristics, including age, education, and income; (ii) labor market characteristics, such as the employment rate, the share of digital labor, and employment shares in sectors affected by government shutdown policies; (iii) health care characteristics, including the location and number of beds of intensive care units (ICUs) and nursing homes; and (iv) territorial and environmental characteristics, such as population density and air pollution, among others. As most of these variables are not available at a regular frequency, we compute their means over the 2015-2019 period and treat them as time-invariant factors. We also source daily data on emergency calls for respiratory reasons and infectious diseases to measure the level of congestion of the emergency care system during the period of the epidemic. Appendix A discusses the data, their sources and coverage in more detail.

## 3 Stylized Facts

Next, we briefly explore four stylized facts that emerge from the data. An in-depth discussion, together with additional supporting figures, is provided in Appendix B. We start by comparing in Figure 1 below daily deaths in 2020 against each of the five preceding years, in the 4,000 municipalities in our sample. Deaths increased rapidly following the detection of Italy’s first community case (denoted by the vertical line). In a month, they more than doubled, from about 800 on February/21^th^ to more than 2,000 on March/21^st^. Our first stylized fact is that COVID-19 may have contributed to the death of more than 24,300 people – almost 0.1% of the local population – from February/21^st^ to March/31^st^ 2020.^2^, ^3^

**Figure 1:**
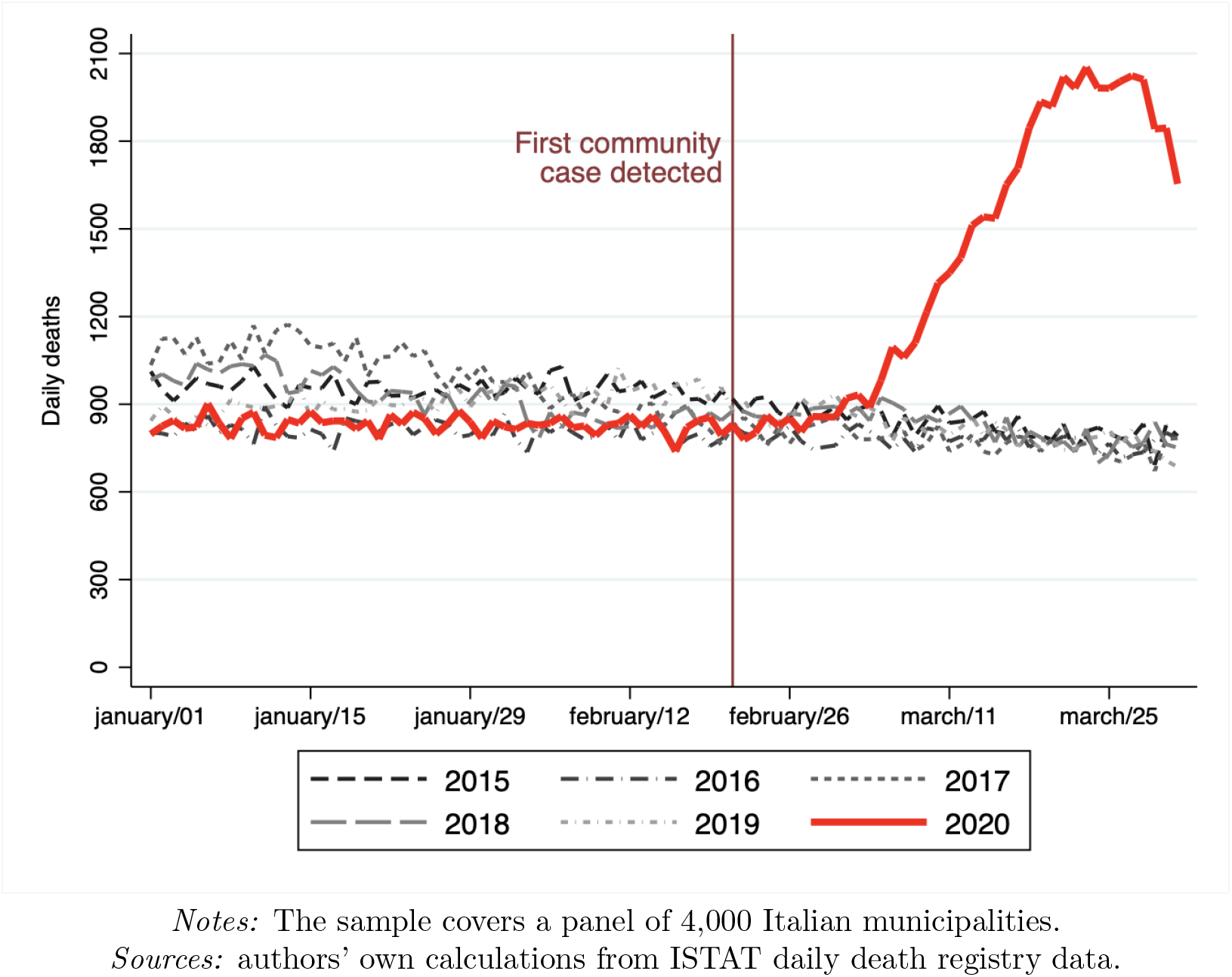
Daily deaths in 2020 compared to the five preceding years.

Next, we compare our estimate of COVID-19 deaths with the number of official COVID-19 fatalities using data from Protezione Civile (2020). This leads to our second stylized fact: COVID-19 deaths have been vastly undercounted in official statistics – plausibly by more than a factor of two (Appendix Figure B1). What can explain this undercounting? In Italy, guidelines for the classification of COVID-19 fatalities vary by region, but in most cases, deaths outside hospitals are not counted in official statistics. Anecdotal evidence suggests that, as the health system struggled with a surge in demand, many old patients may have died of COVID-19 at home or in elderly care facilities (Parodi and Aloisi, 2020), without being counted in official statistics.

To shed more light on the hypothesis above, we explore whether there exist systematic patterns in the rate of undercounting. We find that undercounting was especially high among the elderly and particularly so among women, which is our third stylized fact (Appendix Figure B2). We estimate that, for each official death of a woman in her 80s, two others went undetected. The ratio of undetected to official deaths increases to seven to one when considering women above the age of 90. When we properly account for all these undetected deaths, the gender gap in the number of COVID-19 deaths is sensibly smaller than what it appears in official statistics. According to Italy’s SARS-CoV-2 Surveillance Group (2020), COVID-19 killed more than two men for each woman, while our estimates indicate that deaths among men were only 30% higher than those among women (Appendix Figure B3).

Our fourth and last stylized fact is that COVID-19 had very disproportionate effects across municipalities: in some, the mortality rate increased up to thirty-fold, while in many others, it did not even double. With a few exceptions, the strongest effects of COVID-19 were concentrated in the Lombardia region and some parts of neighboring Emilia-Romagna (Appendix Figure B4).

In the next sections, we formally investigate the insights emerged during this first look of the data. We start with the estimation of the effects of COVID-19 on mortality both across regions and over time.

## 4 The Effect of COVID-19 on the Mortality Rate

To quantify the effect of COVID-19, we need a counterfactual mortality rate in the absence of the virus. We start with a visual inspection of the data and notice that the mortality rate exhibited quite similar trends in 2016 and 2020, up to the day in which Italy’s first community case was detected (see Figure C1 in Appendix C). We then use data from 2015 to 2019 to construct a synthetic control group using the method of Abadie et al. (2010, 2014). This method assigns unit weight to the year 2016, which we thus use as counterfactual.^4^ To estimate the effect of COVID-19 on the mortality rate we rely on a differences-in-differences (DID) approach, in which we normalize the within-year time dimension *t* to take value equal to 0 on February/21^st^ (the day of the first community case), negative values for days before and positive values for those after. For the estimation, we employ the least square method with population analytical weights.^5^ We estimate two specifications, a static and a flexible one. The static specification is as follows:

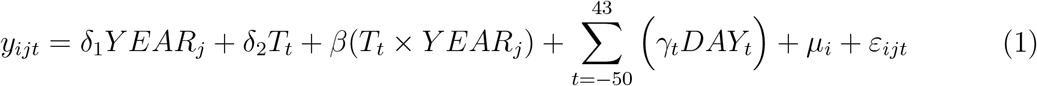

where *y_ijt_* measures daily deaths per 100,000 inhabitants in municipality *i*, at within-year time *t*, for year *j*; *YEAR_j_* is a dummy variable taking value equal to 1 in 2020 and 0 otherwise; *T_t_* is another dummy taking value equal to 1 in the post-February/21^st^ period and 0 otherwise, regardless of the year; *DAY_t_* and *μ_i_* respectively are within-year time and municipality fixed effects; and *ε_ijt_* is an idiosyncratic error, clustered at the municipality-level. The *β* coefficient captures the average effect of COVID-19 on the mortality rate.

For the flexible specification, we replace the pre/post COVID-19 dummy, *T_t_*, with the within-year time fixed effects, *DAY_t_* and estimate the following equation:

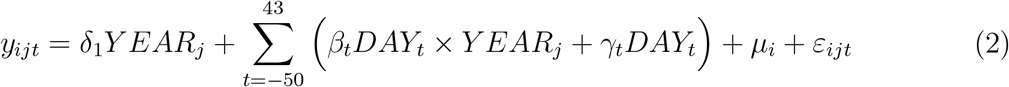

where the notation is as in Equation 1.

We report results in Figure 2 below. Panel A plots the static effect of COVID-19 on daily deaths per 100,000 inhabitants, in the full sample (large red dot) and in each different region (small blue dots), with spikes denoting 95% confidence bands. Panel B plots the effect of COVID-19 on mortality over time, estimated over the full sample of municipalities, with the light blue shaded area denoting the 95% confidence interval.

**Figure 2:**
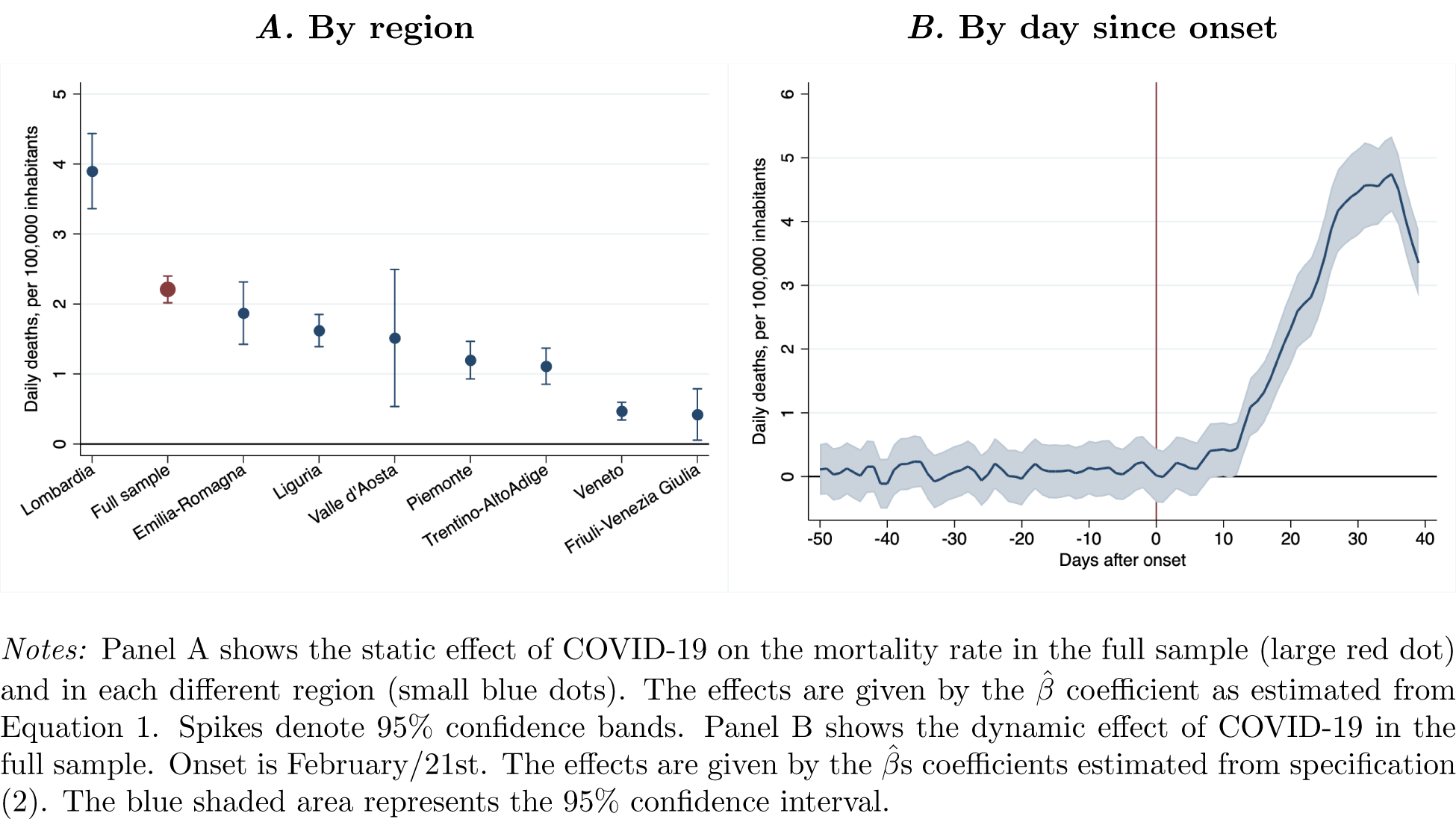
Effect of COVID-19 on daily deaths.

We observe great heterogeneity both across regions and over time. COVID-19 induced the death of slightly more than 2 people per day for each 100,000 inhabitants, on average, during the February/21^st^-March/31^st^ period. This effect was almost twice as large in Lombardia, where the first COVID-19 community case was identified, while it was about four times smaller in Friuli-Venezia Giulia and Veneto. What can explain these differences? The considerable distance between Lombardia and Friuli-Venezia Giulia may explain why the latter was relatively unscathed. Instead, Zanini (2020) and Zingales (2020) attribute the muted effect of COVID-19 in Veneto – which shares a long border with Lombardia – to its different approach to the epidemic management, featuring mass-testing, contact-tracing, and at-home care provision.

Turning to the dynamics, the effect of COVID-19 on mortality peaked 34 days after onset, at about five deaths per day per 100,000 inhabitants, effectively almost tripling the mortality rate (Panel B of Figure 2). The peak was reached 16 days after the government imposed a countrywide lockdown to contain the epidemic, on March/11^th^. Considering an average incubation time of 5.2 days (Lauer et al., 2020; Linton et al., 2020) and a median time of 10 days between the onset of symptoms and death (SARS-CoV-2 Surveillance Group, 2020), the peak timing suggests that the drastic social distancing measures imposed by the government were quite effective in slowing down contagion and – with a lag – mortality.^6^ In the next section, we quantify the effect of the government’s social distancing policies in some more detail.

## 5 The Effects of Shutting Down Businesses

The Italian government took unprecedented measures to fight COVID-19. At the same time in which it imposed a countrywide lockdown, on March/11^th^, it also ordered the closure of all non-essential service activities involving interactions between workers and consumers.

About ten days later, on March/22^nd^, it compounded those measures by ordering the closure of all factories producing non-essential goods.^7^ Many other governments around the world later followed the Italian example and shut down their economies to contain the epidemic. While the social and economic costs of these shutdowns are clear and have been quantified in the literature (see del Rio-Chanona et al., 2020; Koren and Pető, 2020, among others), empirical evidence on their effectiveness in reducing mortality is still scant, which motivates our analysis.

Since we do not have a perfect counterfactual of what mortality would have been in the absence of the government measures, we opt for a ”diff-in-diff” identification strategy a la Rajan and Zingales (1998) and exploit variation in the share of employment in suspended sectors across municipalities. That is arguably exogenous to the government policy since the government ordered the closure of all firms in certain sectors at the national level, rather than deciding the closure of some specific firms in specific municipalities. The identifying assumption is that, if effective, the government measures should have led to larger reductions in mortality in municipalities with a higher share of employment in suspended sectors. This approach allows us only to quantify the differential effect across municipalities of shutting down businesses, but not the overall effect of the policy.

As the government decided to first close down non-essential services (on March/11^th^), and only later it ordered the shutdown of factories (March/22^nd^), we construct two policy interventions dummies. The first takes values 0 and 1 in the pre- and post-March/11^th^ period respectively (*S_t_*), while the second is 1 in the post-March/22^nd^ period and 0 otherwise (*F_t_*). For the estimation, we expand on our static specification (Equation 1) and add the new policy intervention dummies, *S_t_* and *F_t_*, both entering on their own and as an interaction with the year 2020 dummy (*YEAR_j_*) and the employment shares in suspended sectors. The specification that we estimate is as follows:

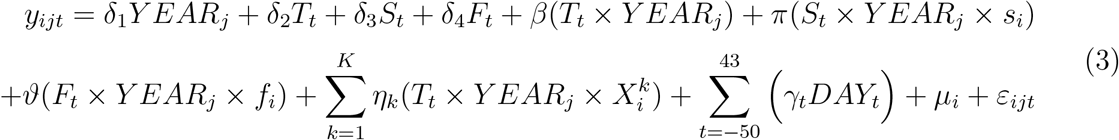

where *s_i_* and *f_i_* are respectively the shares of employment in close down services and factories, in municipality *i*; 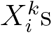 are (time-invariant) municipality characteristics that we include as controls, such as the employment rate, a digitization of work index, an external commuting index and others (see Appendix A); and the rest is as in Equation 1. The *π* and *ϑ* coefficients measure the differential effects, across municipalities, of shutting down non-essential services and factories, respectively.

Table 1 below reports our estimates. We consider both the full sample of municipalities and the restricted sample of Italy’s COVID-19 outbreak epicenter, defined as the area within a radius of 50km from the towns of Codogno or Alzano Lombardo, in the Lombardia region (see Appendix A). We start by estimating a parsimonious specification in which we consider the overall share of employment in suspended sectors, and which does not include any control variable. We then expand on that by, first, distinguishing between services and factories, and, finally, estimating the full specification, also including controls. In the top row, we report the effect of COVID-19 on mortality in the average municipality, while all other coefficients report the effect of an increase of about one standard deviation of the different municipality characteristics (for the employment share in suspended sectors that is 10 percentage points).^8^

When considering the overall share of employment in suspended firms, the estimates are inconclusive (Columns 1 and 4). When we differentiate between sectors, our results suggest that the closure of non-essential service activities was very effective in reducing mortality.

We estimate that the mortality rate was about 15% lower in municipalities with a 10 percentage points higher share of employment in close down services, in both the full and the within-epicenter sample (Columns 2 and 5). Instead, regardless of whether we consider the full sample of municipalities or we only focus on those within the epidemic epicenter, shutting down factories does not seem to have led to significant differences in mortality across municipalities. These results are robust to controlling for many other municipality characteristics that may have had a bearing on COVID-19 mortality, such as external commuting, population density, air pollution and many others (Columns 3 and 6). In Appendix D we also show that the results are very similar when considering a longer time sample, spanning until April/15^th^ and when we control for yet more municipality characteristics.^9^, ^10^

The result that shutting down factories may be less effective in reducing mortality than closing down services squares well with the findings on occupational exposure to COVID-19 of Lewandowski et al. (2020) and Muellbauer and Aron (2020). In particular, the latter uses excess mortality data for England to find that most of COVID-19 deaths in the working-age population were concentrated among people employed in the consumer-facing service sector. While workers in the service sector interact with consumers every day – the opposite of social distancing – for the most part, factory workers only interact with other workers in the same unit, and the opportunities to contract or spread the virus on the workplace appear to be more limited than in the consumer-facing service sector.^11^ Our results also have clear policy implications. Governments should not hesitate in closing down services if they want to reduce COVID-19 mortality. On the other hand, they should carefully weight the less clear benefits of closing down factories against the undoubted costs of the halt in production.

**Table 1:** Suspended firms, municipality characteristics and COVID-19 mortality.

|  | Full sample |  |  | Within epicenter |  |  |
| --- | --- | --- | --- | --- | --- | --- |
|  | (1)<br>Overall<br>suspended | (2)<br>Services vs.<br>factories | (3)<br>With<br>controls | (4)<br>Overall<br>suspended | (5)<br>Services vs.<br>factories | (6)<br>With<br>controls |
| Average effect | 2.25***<br>(0.11) | 2.25***<br>(0.11) | 2.27***<br>(0.11) | 5.98***<br>(0.30) | 5.98***<br>(0.30) | 6.00***<br>(0.28) |
| Suspended firms | 0.00<br>(0.08) |  |  | -0.17<br>0.19 |  |  |
| Suspended services |  | -0.43***<br>(0.09) | -0.34***<br>(0.09) |  | -0.89***<br>(0.26) | -0.80***<br>(0.25) |
| Suspended factories |  | 0.01<br>(0.05) | -0.00<br>(0.04) |  | -0.05<br>(0.11) | -0.06<br>(0.08) |
| Employment rate |  |  | 0.02<br>(0.04) |  |  | 0.50**<br>(0.20) |
| Digital labor |  |  | -0.22**<br>(0.11) |  |  | -1.06***<br>(0.35) |
| External commuting |  |  | 0.36***<br>(0.10) |  |  | 1.10***<br>(0.24) |
| Population density |  |  | 0.72***<br>(0.11) |  |  | 0.17<br>(0.28) |
| Share 80+ |  |  | 0.26**<br>(0.11) |  |  | 2.31***<br>(0.41) |
| High school grads |  |  | -1.17***<br>(0.14) |  |  | -1.85***<br>(0.44) |
| Income inequality |  |  | 0.11<br>(0.10) |  |  | -0.23<br>(0.41) |
| Share health care |  |  | 0.19*<br>(0.10) |  |  | 0.06<br>(0.23) |
| PM10 |  |  | 0.37***<br>(0.10) |  |  | 0.79**<br>(0.38) |
| Observations | 549,256 | 549,256 | 549,256 | 137,923 | 137,923 | 137,923 |
| R-squared | 0.02 | 0.03 | 0.03 | 0.12 | 0.13 | 0.14 |

*Notes:* This table reports the effects of COVID-19 on daily deaths per 100,000 inhabitants by the share of employment in suspended firms. The first row reports the average effect of COVID-19, i.e., $\hat{\beta}$ in Equation (3). Columns 1 and 4 report the effect of having an employment share of 10% in suspended firms, in the full sample and in the epicenter (i.e., 50km radius from Codogno or Alzano Lombardo). Columns 2 and 5 disaggregate the share of suspended firms by services and factories. Columns 3 and 6 extend these results by including a set of controls, also reported in the table. \* \* \* and \*\*\* denote statistical significance at the 90%, 95% and 99% confidence level, respectively.

Before moving on to the next section, we briefly turn to the other variables included in the full specification (Columns 3 and 6). These all enter with the expected sign and help explaining why COVID-19 was so lethal in the Po’ basin (and particularly so in the Lombardia region). This area of Italy has a high population density, with people frequently commuting to other municipalities to work and study, and high levels of air pollution. Our estimates suggest that these are all important risk factors in determining COVID-19 mortality.^12^

Among mitigating factors, a higher share of digital labor is associated with lower COVID-19 mortality, suggesting that smart working policies may have been effective in reducing COVID-19 mortality. But it is education to have the strongest mitigating effects. A 7.5 percentage point higher share of high school graduates is associated with 20% to 50% lower COVID-19 mortality. What might explain this result? Education is usually associated with better underlining health conditions (Case and Deaton, 2017; Chetty et al., 2016) and co-morbidities are one of the main COVID-19 risk factors (Yang et al., 2020). Another, nonexclusive, possibility is that more educated people may be very conscious about the risks associated with COVID-19 and thus more pro-actively engage in social-distancing practices (Adda, 2016; Wright et al., 2020).

## 6 Health Care Management within the Epicenter

We next zoom in on Italy’s COVID-19 outbreak epicenter – including municipalities within a radius of 50km from the towns of Codogno or Alzano Lombardo, in the Lombardia region (see Appendix A for details). This is the most densely populated and among the wealthiest areas of Italy. With almost 1,000 municipalities, it makes up for about 10% of the country’s total population and more than 15% of its national income. It is also renowned for having one of the best health care infrastructures in the country, which itself has relatively high rates of intensive care units (ICUs) per capita (McCarthy, 2020). However, the force with which the virus struck was so intense that it brought the system close to collapse. Using our flexible specification (Equation 2), we estimate that COVID-19 mortality reached a peak of 12 people per day per 100,000 inhabitants, which is four times more than outside the epicenter (see Figure E1 in Appendix E).

There is ample anecdotal evidence that the health care system got overwhelmed (see, for instance, Beall, 2020; Johnson, 2020; Sorbi, 2020). In this section, we quantify by how much this may have contributed to the high COVID-19 mortality. This is key to understand how better preparedness could help to save lives when new outbreaks materialize. We consider, in particular, the role played by the emergency transportation system. As highlighted by SARS-CoV-2 Surveillance Group (2020) and Sun et al. (2020) among others, getting intensive care on time is key for the survival of critical COVID-19 patients. A longer ambulance ride to the ICU may thus make a difference in the survival probability. In what follows, we test whether COVID-19 mortality increases with distance from the ICU. The focus is on the Lombardia region, which accounts for 95% of all municipalities in the outbreak epicenter, and for which we have good health care data.

We start by sourcing data on location and areas of specialization of each hospital and private clinic and construct a municipality-level variable measuring distance to the nearest ICU. If there is an ICU in town, we set it to zero. Otherwise, the variable measures the linear distance to the nearest municipality with an ICU, in km (see Appendix A for more details).^13^ A first look at this variable reveals that municipalities with an ICU in town are less than 4% and account for just over 25% of the population. For more than 40% of all municipalities, the closest ICU is further than 10 km, meaning that transportation there may involve a significant trip.^14^

Of course, distance on its own does not imply that critical patients cannot get to the ICU on time. After all, also in normal times some patients require transportation from distant municipalities to the ICU, and usually they get there on time. But during the COVID-19 epidemic, the burden on the emergency care system was so high that it might have forced emergency staff to prioritize serving more patients at the expense of reducing geographical coverage (see Vergano et al., 2020, for evidence on the existence of prioritization guidelines). In Figure E2 in Appendix E we show evidence that, as COVID-19 was making inroads among the population, calls to the emergency line soared, increasing up to five times more than their level during normal times. As the system became overwhelmed, waiting times for emergency transportation swelled. Sorbi (2020) reports that to make a trip that usually took only 8 minutes, ambulances were taking an hour, and in some cases, they were not getting in on time.

To test whether the congestion of the emergency system may have resulted in higher mortality rates in municipalities far from the ICU, we proceed in two steps. We first assess whether municipalities with an ICU experienced a lower COVID-19 mortality during the epidemic. We then test whether this effect was larger during periods of emergency care congestion, which we measure using the daily volume of calls to the emergency system for respiratory reasons or infectious diseases (see van Dijk et al., 2008, for evidence on the volume of emergency calls as a predictor of emergency transportation availability).

We start by expanding Equation (1) to estimate a specification in which we interact distance to ICU to the COVID-19 treatment variable, as follows:

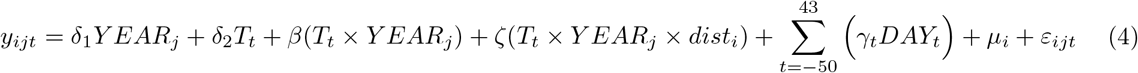

where *dist_i_* is distance to ICU for municipality *i* and the rest is as in Equation 1. We run the estimation separately by gender and age, which allows us to also explore interactions between COVID-19 and demographics. In Panels A and B of Figure 3, we show the effect of COVID-19 on mortality in municipalities with an ICU in town (crosses) and in those where the closest ICU is at 10 Km (dots), for respectively the working age people and the elderly.^15^ We then estimate a dynamic specification to evaluate the effect of distance to ICU over time (similarly as above, but using the within-year effects as treatment, see Equation 2). Figure 4 compares the dynamic effect of distance from the ICU, on average across all ages, to the volume of daily calls to the emergency system for respiratory reasons and infectious diseases. The figures below are for men. Results for women are aligned and are shown in Appendix Figure E.

**Figure 3:**
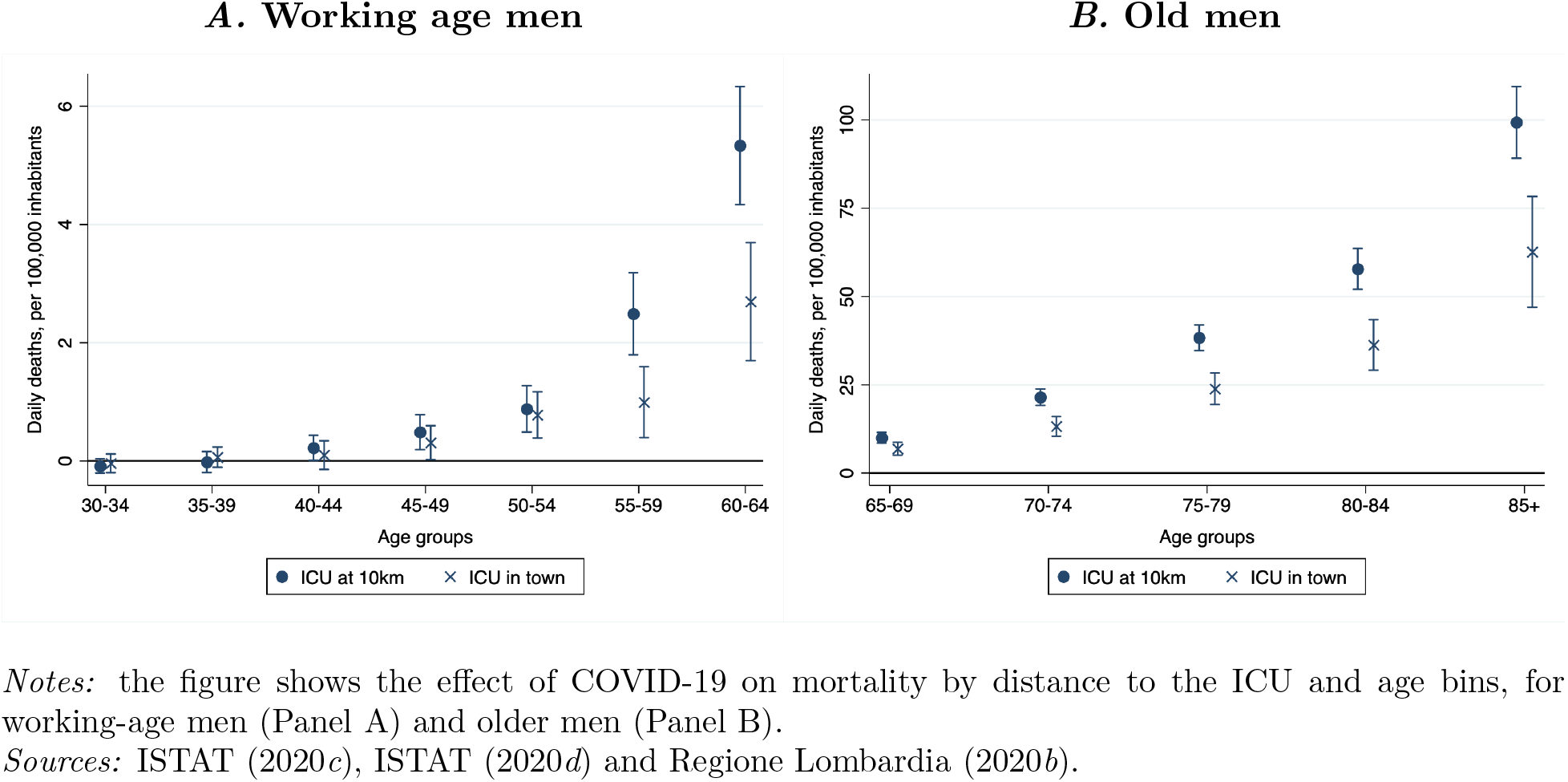
Distance to ICU and COVID-19 mortality, by age.

Strikingly, we find that COVID-19 had a significant effect on mortality on men as young as 40 years old (Figure 3, Panel A). Among men above 55, municipalities with an ICU in town consistently experienced 30% to 50% lower mortality than those distant 10km from an ICU (Panel B). Looking at the dynamics, the additional effect on mortality of being far from the ICU became larger as the number of incoming emergency calls swelled – a sign that the congestion of the emergency care system may have prevented critical patients from being transported to the ICU on time. At the peak, municipalities distant 10km from the ICU experienced 6 more deaths per day per 100,000 inhabitants than municipalities with an ICU in town – almost twice as much (Figure 4). In Table E1 in Appendix E, we show that these results are robust to controlling for other municipality characteristics that may correlate with distance to ICU and that could also have an effect on mortality, such as population density, external commuting, the share of high school graduates, that of health care workers and many others. We also check that the effect of distance to the ICU on mortality is not observed outside the outbreak epicenter, another sign that distance to ICU only matters when the system is overburdened.

**Figure 4:**
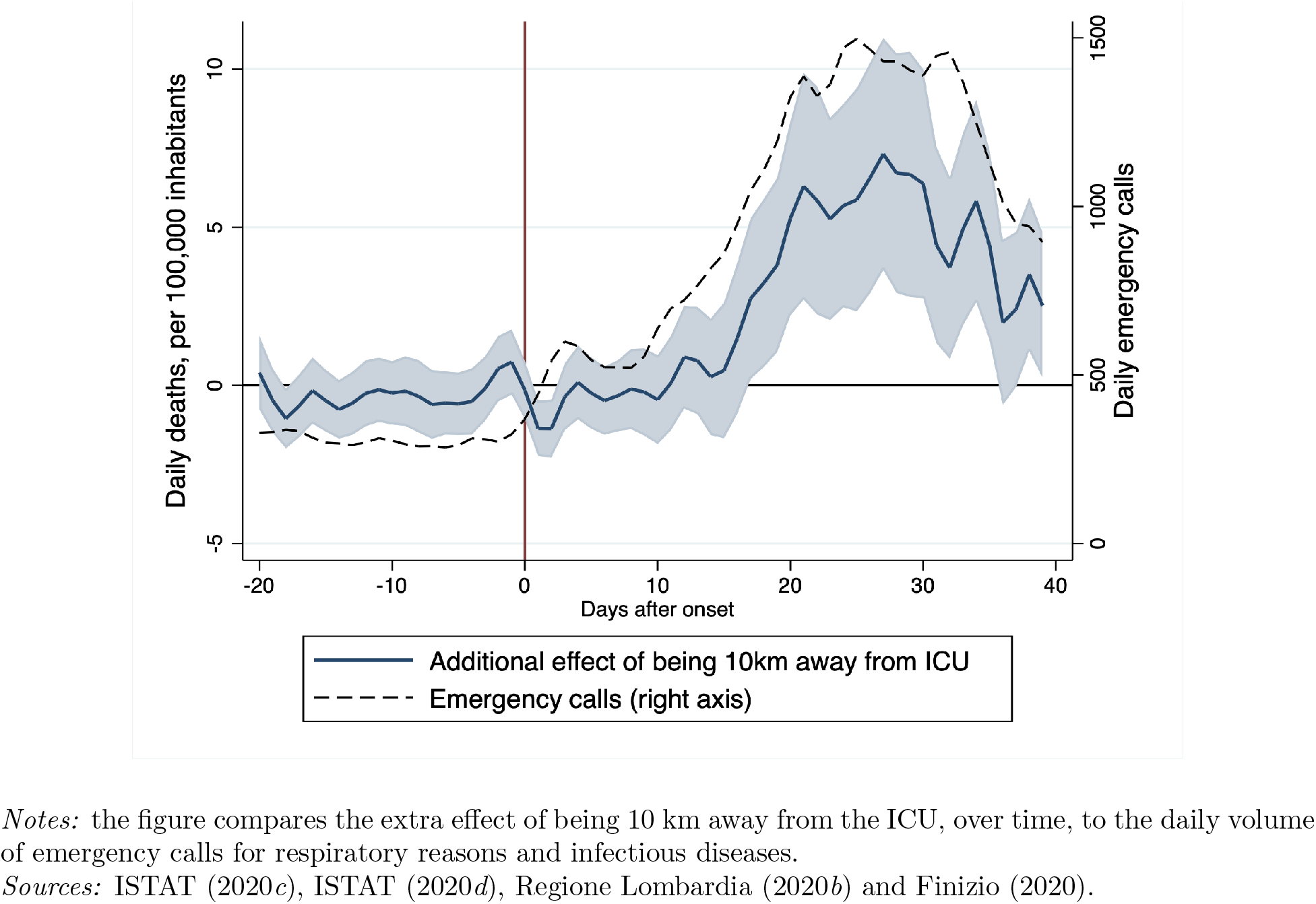
Distance to ICU and COVID-19 mortality at times of system congestion.

Our results suggest that many COVID-19 deaths may have been prevented through better preparedness. Drawing a lesson from Italy’s tale, governments around the world should invest in strengthening their emergency care response. They should improve prehospital emergency services, by clarifying the first point of contact for possible COVID-19 cases, expanding capacity to manage large volumes of calls, and improving phone triage to better prioritize care delivery. They should also invest in building ambulance capacity, and, ideally, mobilizing ICUs more evenly across the territory. All these factors would be key to help reduce mortality shall new outbreaks materialize.

In Appendix F we discuss some additional analysis that we carried out regarding the management of the health care sector in Italy’s COVID-19 outbreak epicenter, specifically focusing on nursing homes. These have been in the spotlight across Europe and beyond for being possible hotspots of contagion and deaths. Our results for Italy suggest that living in a nursing home may have significantly increased the probability of dying during the COVID-19 epidemic. We find that in municipalities with a 10 percentage point higher share of people living in a nursing home, mortality among old men was about 30% higher than in municipalities without a care home. The results are even more striking for women – mortality was about 50% higher very old women – and help rationalizing the serious undercounting of COVID-19 fatalities in official statistics (discussed in Section 3), which do not include nursing home deaths.

## 7 Conclusion

In this paper we zoomed in on two key policy aspects of the COVID-19 epidemic – business shutdowns and the management of the emergency care system. The first aims at reducing the pace and extent of COVID-19 infections, while the second is crucial to ensure that everyone in need receives timely care. Both of them can dramatically affect the human and economic toll of COVID-19. Our study provides useful lessons for policymakers shall new outbreaks materialize.

The analysis draws from highly granular death registry data for a sample of 4,000 Italian municipalities, as well as detailed health care data and data on employment in sectors shut down by the government. To assess the effectiveness of business shutdowns, we exploited exogenous variation across municipalities in the employment shares in shut down businesses. Our results, which are robust to controlling for a host of co-factors, offer strong evidence that the closure of service activities is very effective in reducing COVID-19 mortality. We find that mortality was about 15% lower in municipalities with a 10 percentage points higher employment share in shut down services. On the other hand, our results suggest that shutting down factories is much less effective. We hypothesize that this is because factory workers engage in limited physical interactions relative to workers in the consumer-facing service sector. These results have clear policy implications. In mitigating COVID-19, governments should not hesitate to close down services, but they should more carefully weight the less clear benefits of closing down factories against the undoubted costs given by the halt in production.

Concerning the management of the health care system, we find that mortality strongly increases with distance from the intensive care unit (ICU). Municipalities at 10 km from the closest ICU experienced up to 50% higher mortality. This effect was strongest within the epicenter and in days with abnormally high volumes of calls to the emergency line. Our results highlight the importance of increasing preparedness, both in Italy and abroad, to help reduce mortality shall new outbreaks materialize. Governments should improve pre-hospital emergency services, build ambulance capacity, and, ideally, mobilize ICUs more evenly across the territory.

## Data Availability

Data available upon request

## Appendix

### A Dataset

This Appendix discusses the data in more detail. Death registry data are taken from ISTAT (2020 c). Following the COVID-19 outbreak, ISTAT (2020*c*) started disseminating death registry data for a subsample of the municipalities reporting to the national registry of the resident population (ANPR) – which are about three-quarters of Italy’s total. Initially, ISTAT (2020*c*) only reported data for the subsample of municipalities that were deemed to provide accurate information and had experienced an increase in mortality in 2020 of at least 20% relative to the average of the five preceding years. In early May 2020, ISTAT (2020c) released data for the January/1^st^-March/31^st^ period for all the municipalities which had provided accurate information – regardless of whether they report to ANPR or had experienced an increase in mortality of more than 20%. Only 9% of municipalities in the eight regions we focus on, accounting for about 7% of the population, were excluded. At the same time, in early May 2020, ISTAT (2020c) also released data for the first 15 days of April, for the sample of municipalities that reported to ANPR. Our main focus is on the larger sample of municipalities (thus only covering the January/1^st^-March/31^st^ period). We use the remaining data to perform some extensions. Table A1 below reports sample statistics.

Regarding socio-demographic characteristics, COVID-19 is thought to be more lethal among men and the elderly (SARS-CoV-2 Surveillance Group, 2020). We thus consider the share of those aged 80 and above and the share of women in the population. We also consider the share of high-school graduates over the working-age population and a variable measuring income inequalities (the ratio of the total income of the richest 20% to the total income of the poorest %). Education and income may affect the lethality of COVID-19 through two main channels. For one, low-income agents tend to have worse health (Case and Deaton, 2017; Chetty et al., 2016), and Yang et al. (2020) have shown that patients with co-morbidities have a higher chance of dying from COVID-19.^16^ Second, income and education are also likely to affect health behavior (see Galama et al., 2018) and attitudes towards, and the feasibility of, social-distancing practices (Adda, 2016). We also collect data on inhabited land and construct a population density variable.

**Table A1:**
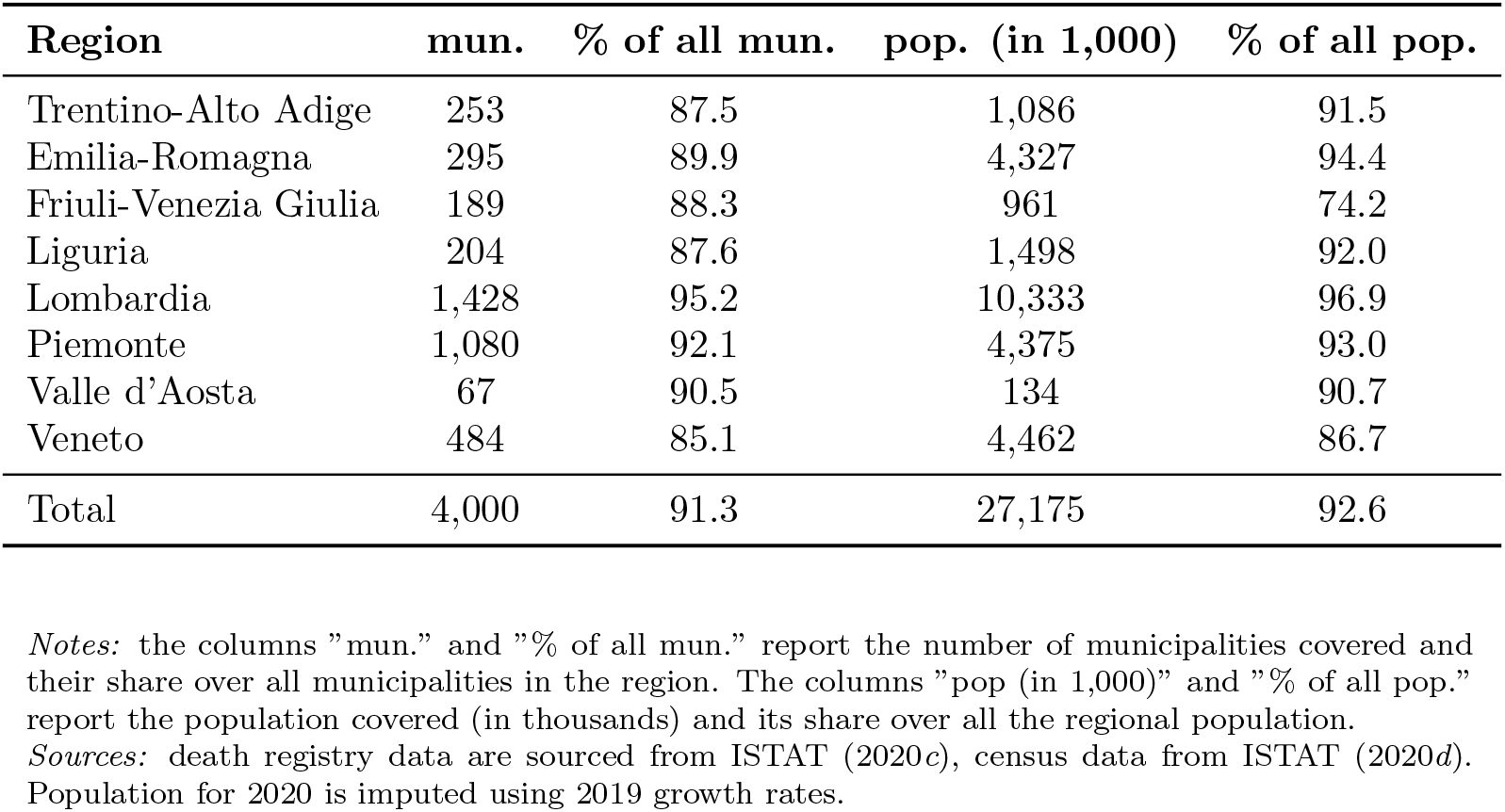
Population coverage of death registry data

| Region | mun. | % of all mun. | pop. (in 1,000) | % of all pop. |
| --- | --- | --- | --- | --- |
| Trentino-Alto Adige | 253 | 87.5 | 1,086 | 91.5 |
| Emilia-Romagna | 295 | 89.9 | 4,327 | 94.4 |
| Friuli-Venezia Giulia | 189 | 88.3 | 961 | 74.2 |
| Liguria | 204 | 87.6 | 1,498 | 92.0 |
| Lombardia | 1,428 | 95.2 | 10,333 | 96.9 |
| Piemonte | 1,080 | 92.1 | 4,375 | 93.0 |
| Valle d'Aosta | 67 | 90.5 | 134 | 90.7 |
| Veneto | 484 | 85.1 | 4,462 | 86.7 |
| Total | 4,000 | 91.3 | 27,175 | 92.6 |

*Notes:* the columns "mun." and "% of all mun." report the number of municipalities covered and their share over all municipalities in the region. The columns "pop (in 1,000)" and "% of all pop." report the population covered (in thousands) and its share over all the regional population.
*Sources:* death registry data are sourced from ISTAT (2020c), census data from ISTAT (2020d). Population for 2020 is imputed using 2019 growth rates.

Moving on to labor market characteristics, these may have an important role in the diffusion of COVID-19. A higher level of economic activity is likely to increase physical contacts, thus leading to higher contagion. Along these lines, Markowitz et al. (2019) show that higher employment rates are associated with a higher flu incidence. Hence, we consider the employment rate as a potential factor affecting mortality. Other labor market characteristics may also play a role. We expect contagion to be higher in areas where a lower share of jobs can be done digitally, which should thus increase mortality. We construct a continuous 0-1 digitalization of work index, which assigns higher values to municipalities where a higher share of jobs is/can be done digitally.^17^ We also source an index measuring the level of external commuting, that is the level of inflows into a municipality for work and study reasons (from other municipalities). As shown in Adda (2016); Harris (2020), commuting is an important determinant of contagion. Finally, we note that the Italian government ordered the closure of all non-essential consumer-facing service activities on March/10^th^ and that of all construction and manufacturing industries producing non-essential goods on March/17^th^. We thus construct three variables measuring the share of employment in (i) overall suspended industries, (ii) suspended industries in services, and (iii) suspended industries in manufacturing. For that, we use data on overall employment from ISTAT (2020a) and on suspended sectors from ISTAT (2020b).

As for health care characteristics, we focus on the share of employment in the health care sector, the availability of intensive care units (ICUs), and the reliance on nursing homes to provide for the elderly. As health care workers are the most exposed, we expect the share of employment in the health care sector to be correlated with higher contagion and mortality. As for ICU, SARS-CoV-2 Surveillance Group (2020) analyze individual-level data on all official COVID-19 deaths in Italy and find that the usage of ICUs has doubled the average time between hospitalization and death, suggesting that it may have also saved lives. Unfortunately, ICUs are not available in every municipality, and some hospitals became overburdened with COVID-19 patients, to the point that they could not provide ICUs to everyone in need (see reports by Beall, 2020; Vergano et al., 2020). We construct a variable measuring distance (in km) to the closest ICU, using data on public and private health care facilities collected from Regione Lombardia (2020*b*) and geospatial coordinates (ISTAT, 2020*e*). When there is an ICU in a municipality, distance is set to zero.^18^ Nursing homes have been in the spotlight for being a catalyst of infections and deaths among the elderly, due to close physical proximity among staff and residents and lack of preparedness (The Economist, 2020*b*). We retrieve data on the number of beds available in each facility to construct two variables. The first variable measures the distance from the closest nursing home. The second, captures the total number of available places in nursing homes in each municipality where there is at least one. Relevant data come from Regione Lombardia (2020*a*) and ISTAT (2020*e*). The variables for ICUs and nursing homes only cover the Lombardia region. To calculate the measure of the proportion of elderly in nursing homes, we use information on the age and sex composition of nursing home patients that we multiply with the number of beds in each municipality. Under the assumption of full occupancy, this gives us the number of beds occupied by each age group and sex.^19^ We then divide the number of beds per age group and sex by the corresponding population.

As for environmental and territorial characteristics, we focus on air pollution and distance from the epicenters, which we identify as the two towns of Codogno and Alzano Lombardo (see the discussion in Appendix B below). Several studies have shown long-term exposure to particulate matters such as PM10 and PM2.5 to increase health risks (for a review, see World Health Organization, 2003), and Wu et al. (2020); Conticini et al. (2020); Becchetti et al. (2020) have found a positive link between both PM10 and COVID-19-induced mortality (the latter for a sample of Italian provinces). Since data availability on air pollution at the municipality is an issue, we use data on the number of days in a year in which the level of PM10 is above the limit, in each province, and match it to municipalities.^20^ As for distance from the epicenter, we construct a variable measuring the minimum distance (in km) from either Alzano Lombardo or Codogno. We define municipalities inside the epicenter as these municipalities within a radius of 50km from either of the two towns.

We also collect additional variables, such as the before-tax mean income, the share of the Chinese immigrants in the population, an index measuring the quality of sanitary facilities in houses, an index measuring internal commuting, and the share of working-age people with a temporary employment contract.

Since all the variables capturing municipality characteristics are available at an irregular frequency, we compute their mean over the 2015-2019 period and treat them as time-invariant characteristics. We also censor all observations that are four standard deviations above and below the median. A list of all variables, their source, and descriptive statistics are provided in Table A2 below.

**Table A2:**
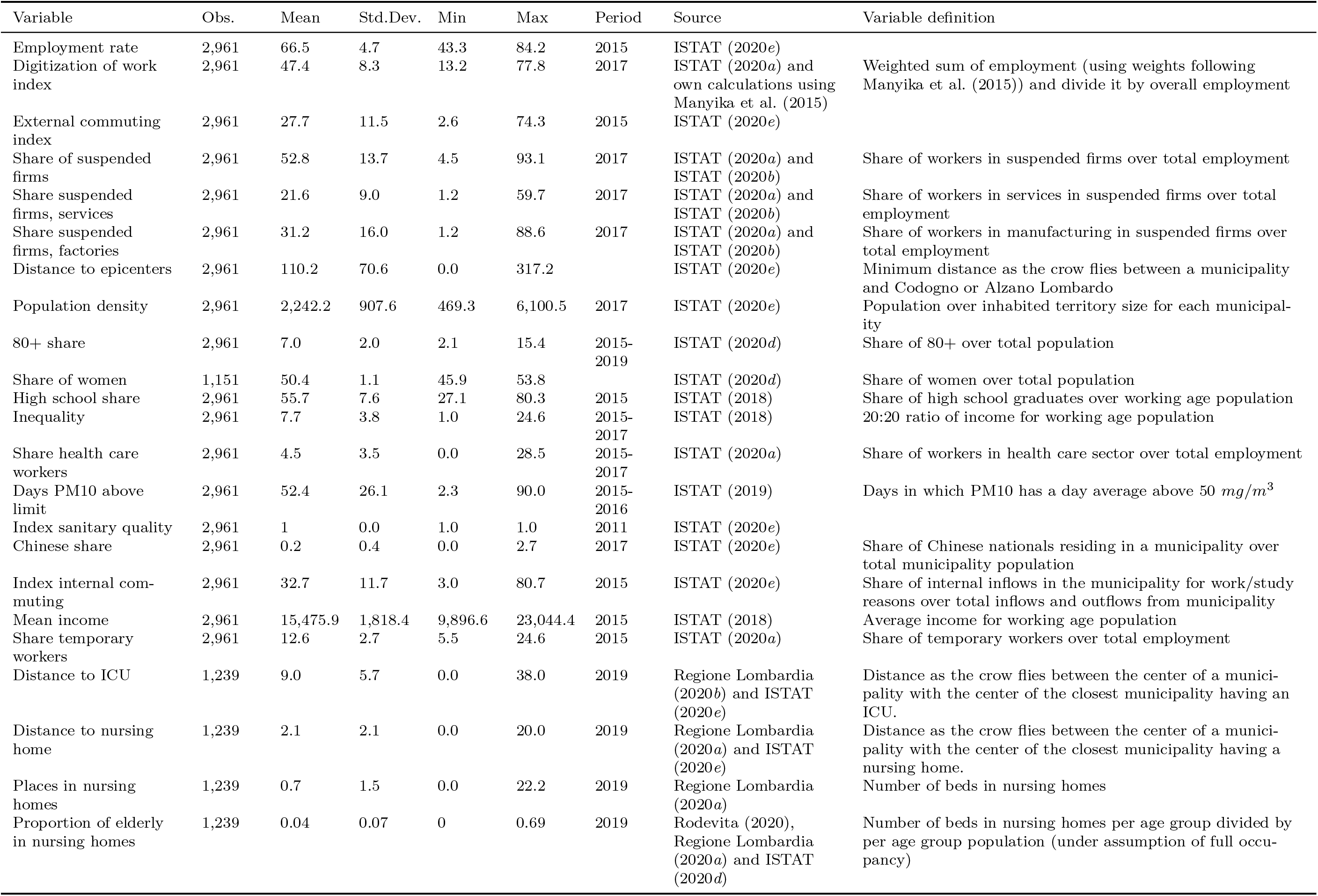
Variable descriptives, source and definitions

### B Stylized Facts

This appendix complements the discussion on graphical evidence of Section 2 and provides relevant figures. Figure B1 compares excess deaths from death registry data to official COVID-19 fatalities from Protezione Civile (2020).

**Figure B1:**
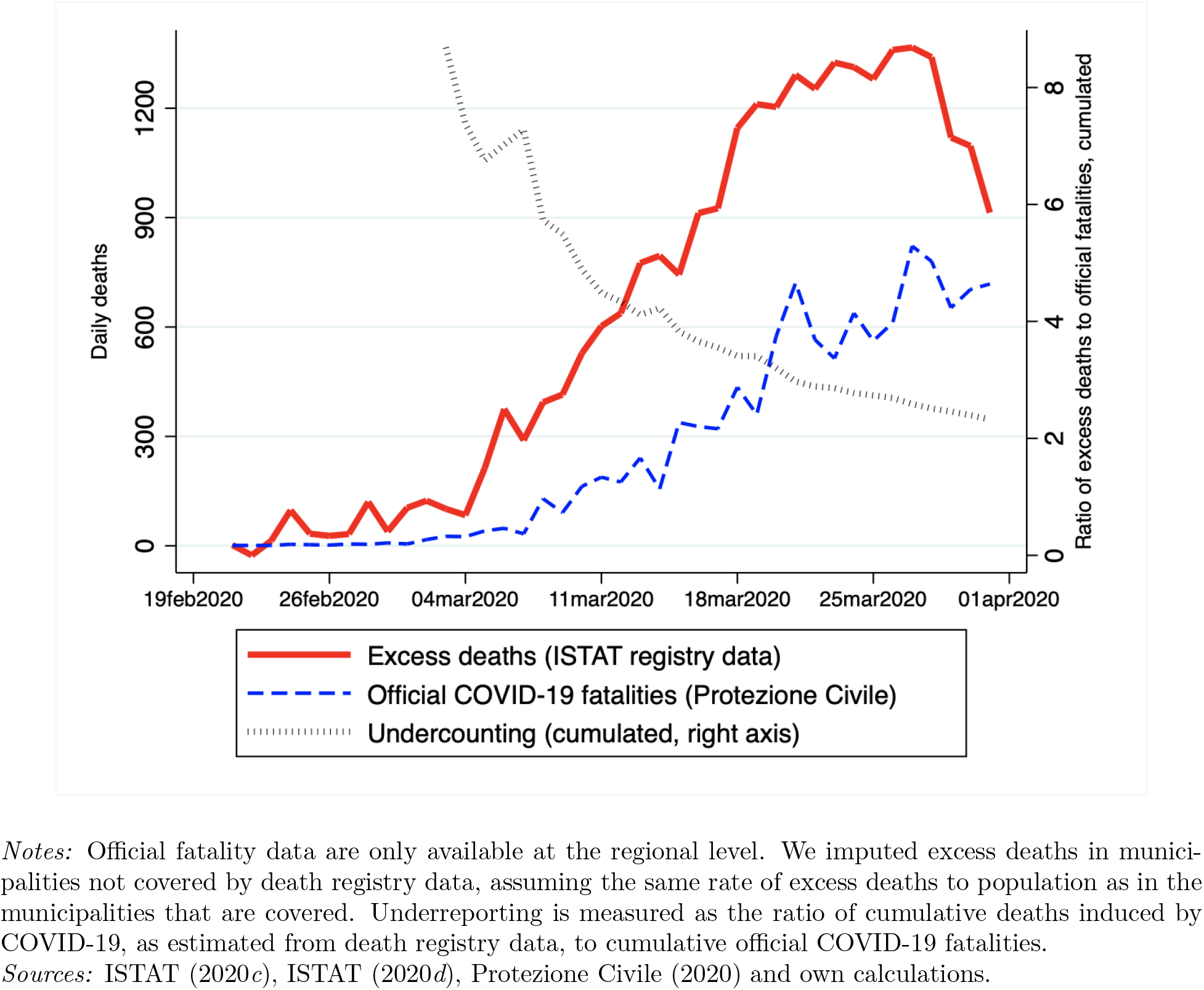
COVID-19 deaths from registry data compared to official statistics

As of March/31^st^, the ratio of excess deaths from registry data to official COVID-19 fatalities is 2.3. As discussed in Footnote 3, excess deaths also include people who died without having contracted COVID-19 (indirect deaths). To gauge the importance of indirect deaths, we consider the 12 regions of Italy which are not included in our sample because they were relatively unaffected by COVID-19. These display ratios of excess deaths to official COVID-19 fatalities that are similar to the regions in our sample (about 2,800 excess deaths to 1,400 official COVID-19 fatalities, overall). Assuming that these regions correctly detected all COVID-19 deaths, we obtain a measure of indirect deaths as a share of the population, which we use to infer the number of indirect deaths in the eight regions in our sample. We then subtract this number from the number of excess deaths and obtain a measure of COVID-19 deaths (direct deaths). Comparing the number of direct deaths against official fatalities, we estimate a rate of undercounting of about 2.2 – meaning that for each reported death, an additional 1.2 may have gone undetected.

The scale of undercounting was very high at the beginning of the epidemic and progressively decreased. This suggests that limited testing capacity may be a potential reason for undercounting, as authorities may have initially lacked the necessary testing capacity to detect all cases.

**Figure B2:**
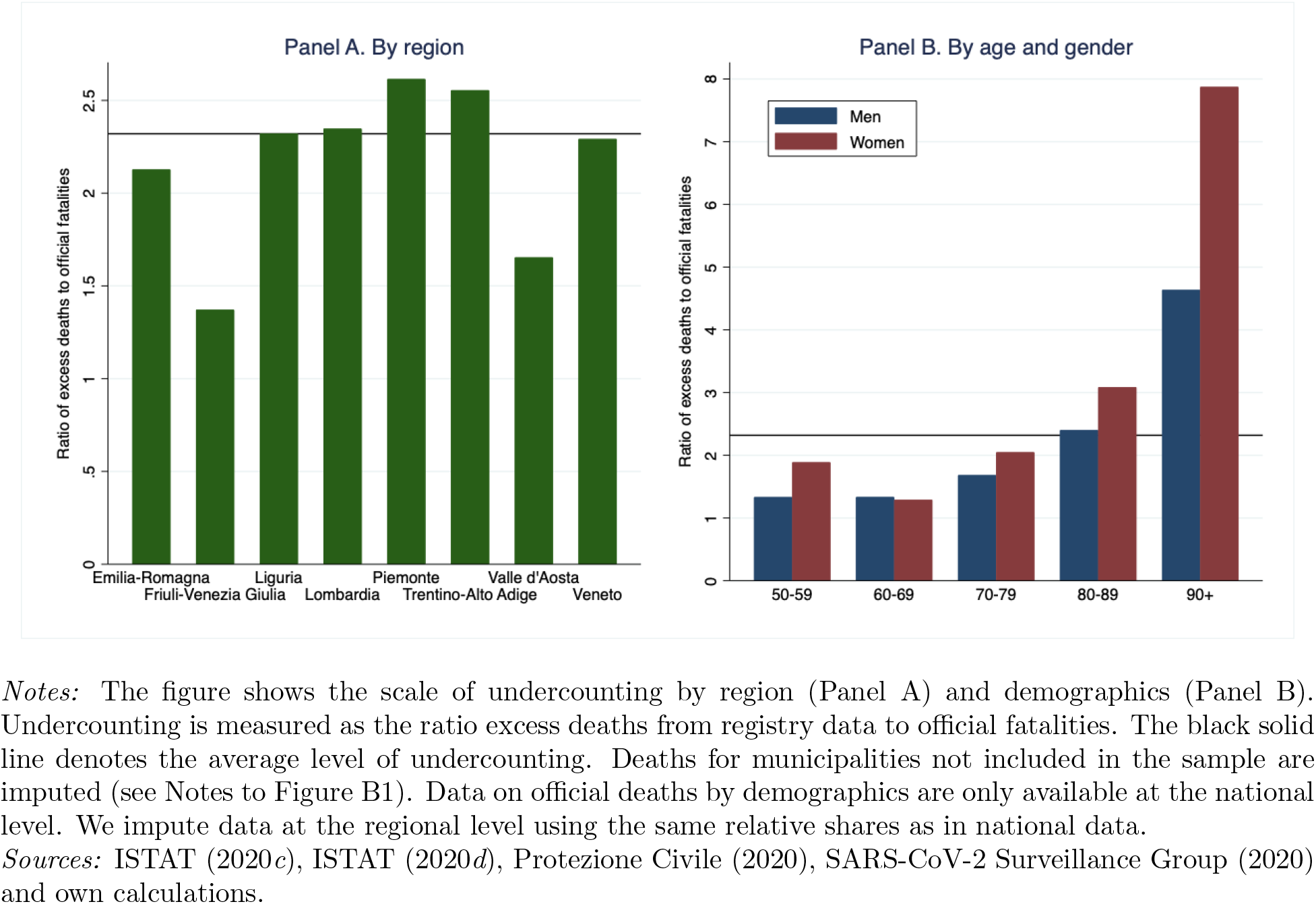
Undercounting of COVID-19 deaths in official statistics

Figure B2 above shows the scale of undercounting by regions (Panel A) and demographics (Panel B), while Figure B3 below compares total excess deaths from death registry data against COVID-19 official fatalities, by age and gender. The figures show that the extent of undercounting is particularly high among older women. After accounting for all the deaths of older women that went undetected, the gender gap in the number of deaths caused by COVID-19 is sensibly reduced. According to official statistics, COVID-19 killed more than two men for each woman, while our estimates indicate that deaths among men were only 30% higher than those among women.

**Figure B3:**
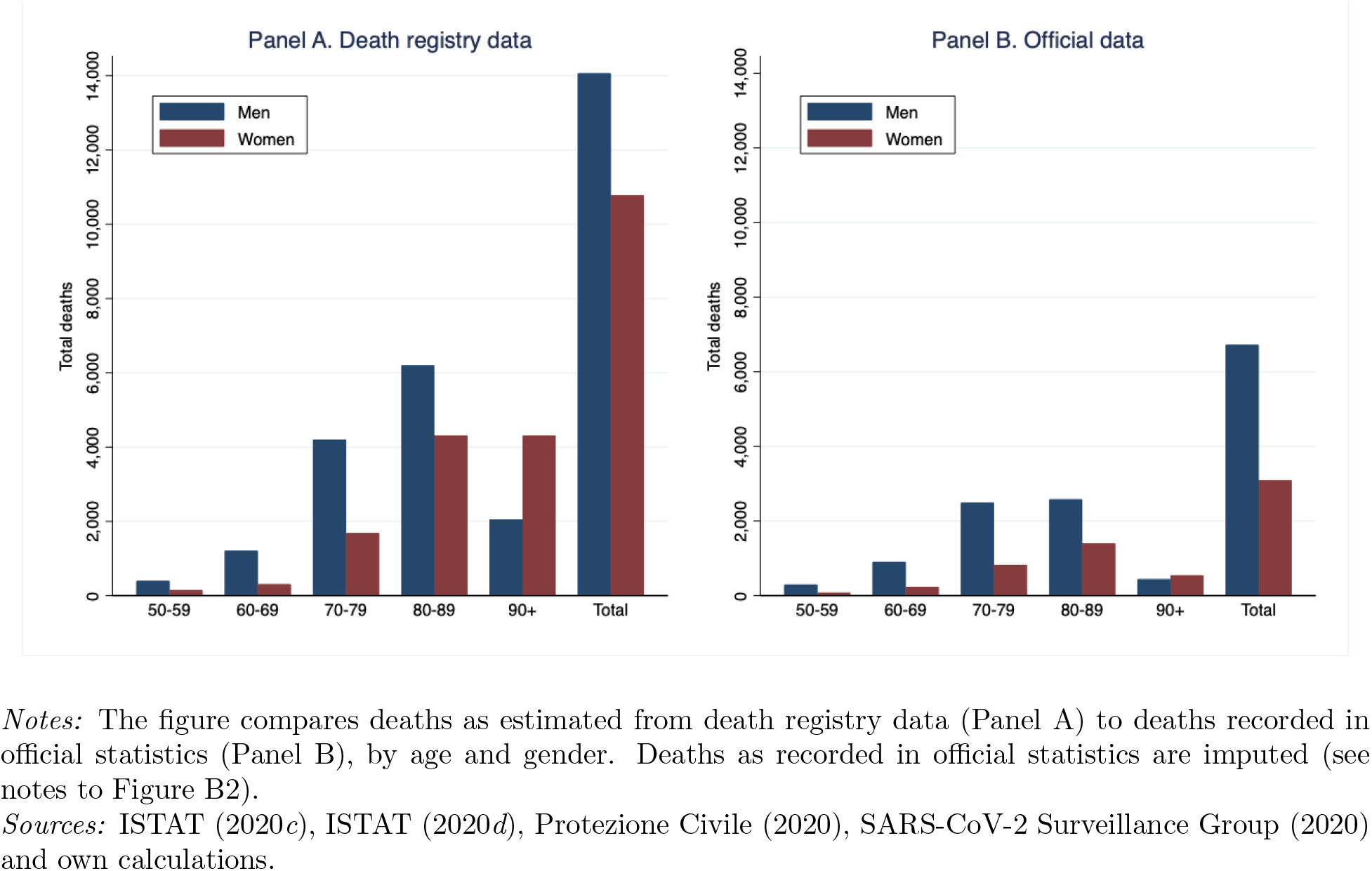
COVID-19 deaths by age and gender

We next move on to daily mortality rates per 100,000 inhabitants and explore variation in mortality across the 4,000 municipalities covered in the dataset. Figure B4 below depicts the extent of the epidemic both over time and across space. Two large outbreaks are apparent in the Lombardia region, one in the south, around the town of Codogno, where the first community case was detected, on February/21^st^, and another in the north, around Alzano Lombardo, in the Bergamo province. Some reports argue that the virus was likely circulating among patients admitted at the local hospital in Alzano Lombardo already before the first community case was detected on February/21^st^ (see, for instance, Imariso and Ravizza, 2020)). On this basis, we consider the area within a radius of 50km from the towns of Codogno or Alzano Lombardo as the epicenter of Italy’s COVID-19 outbreak.

**Figure B4:**
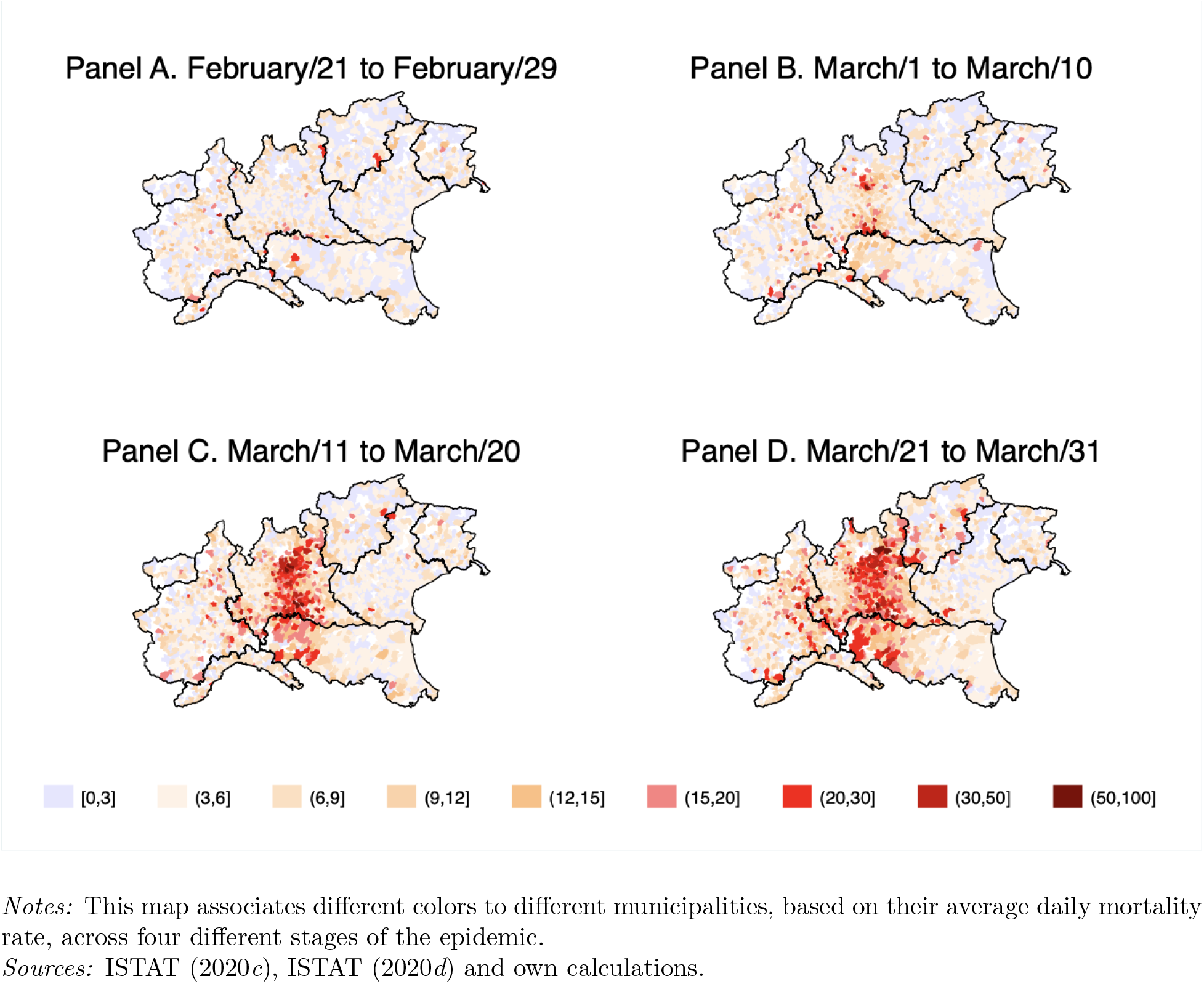
Mortality Rate Across Municipalities and Over Time

### C Additional Results and Robustness Checks on Main Estimation

**Figure C1:**
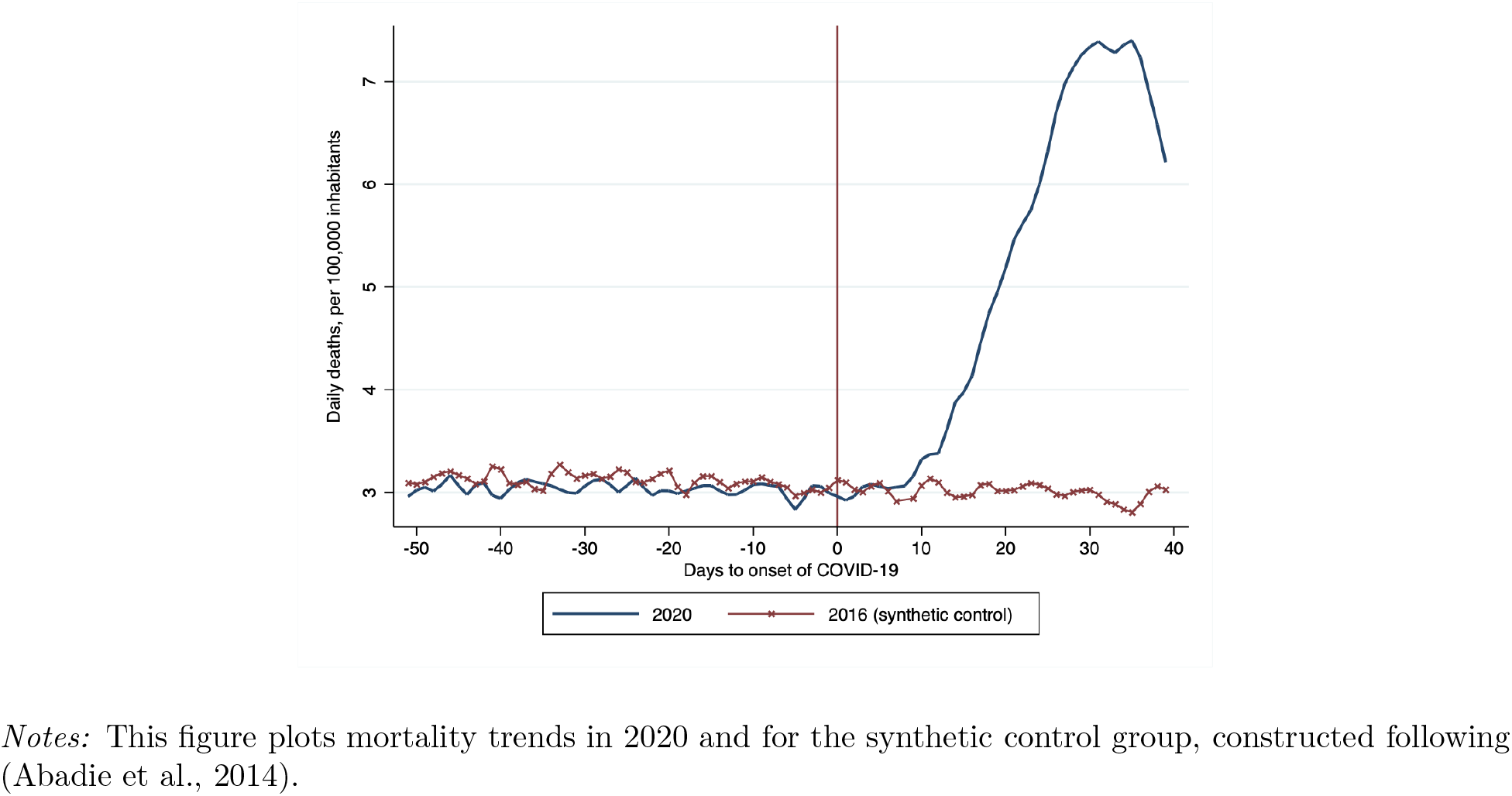
Daily mortality trends

**Figure C2:**
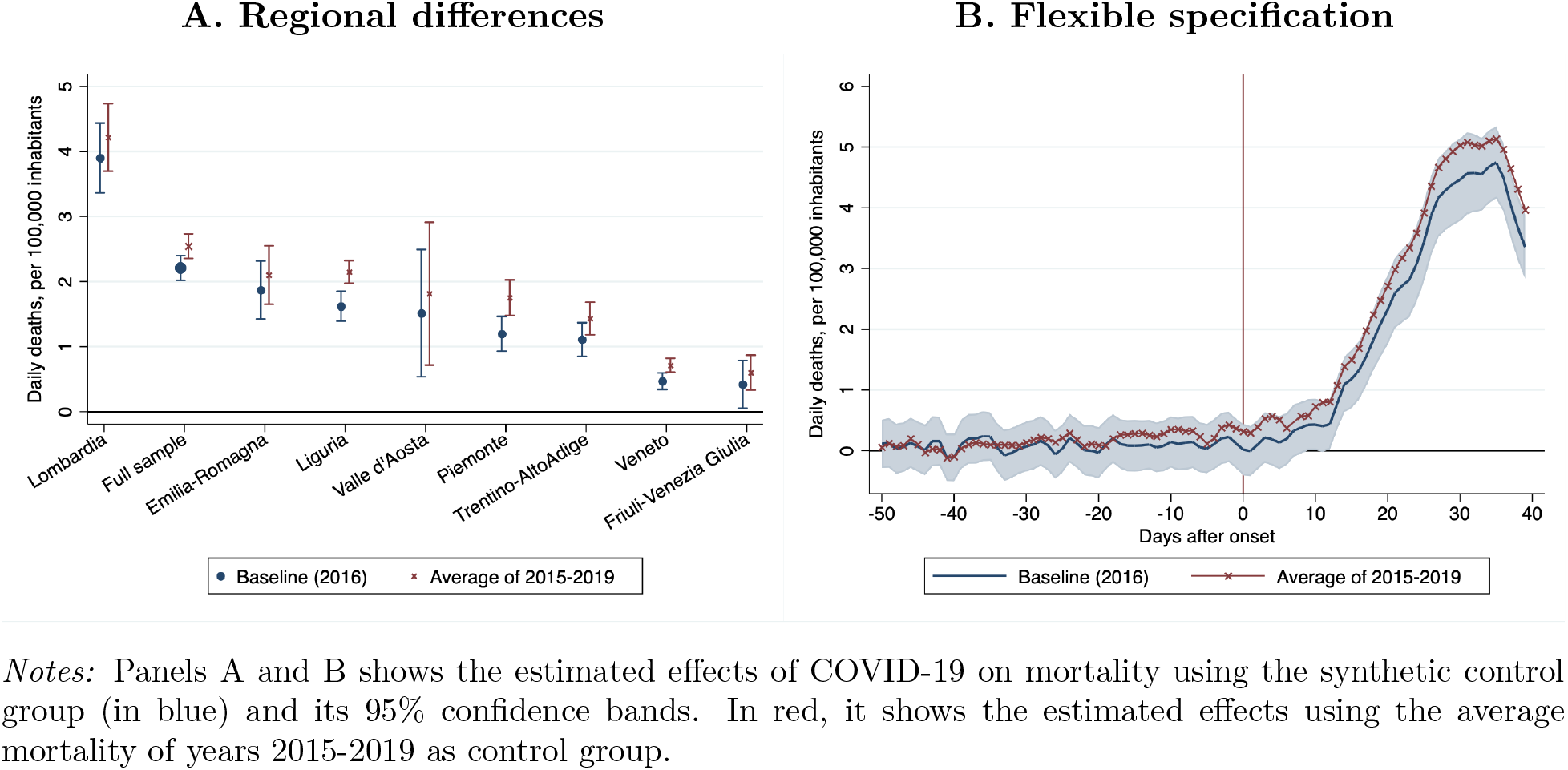
Robustness of the estimates to the control group

**Figure C3:**
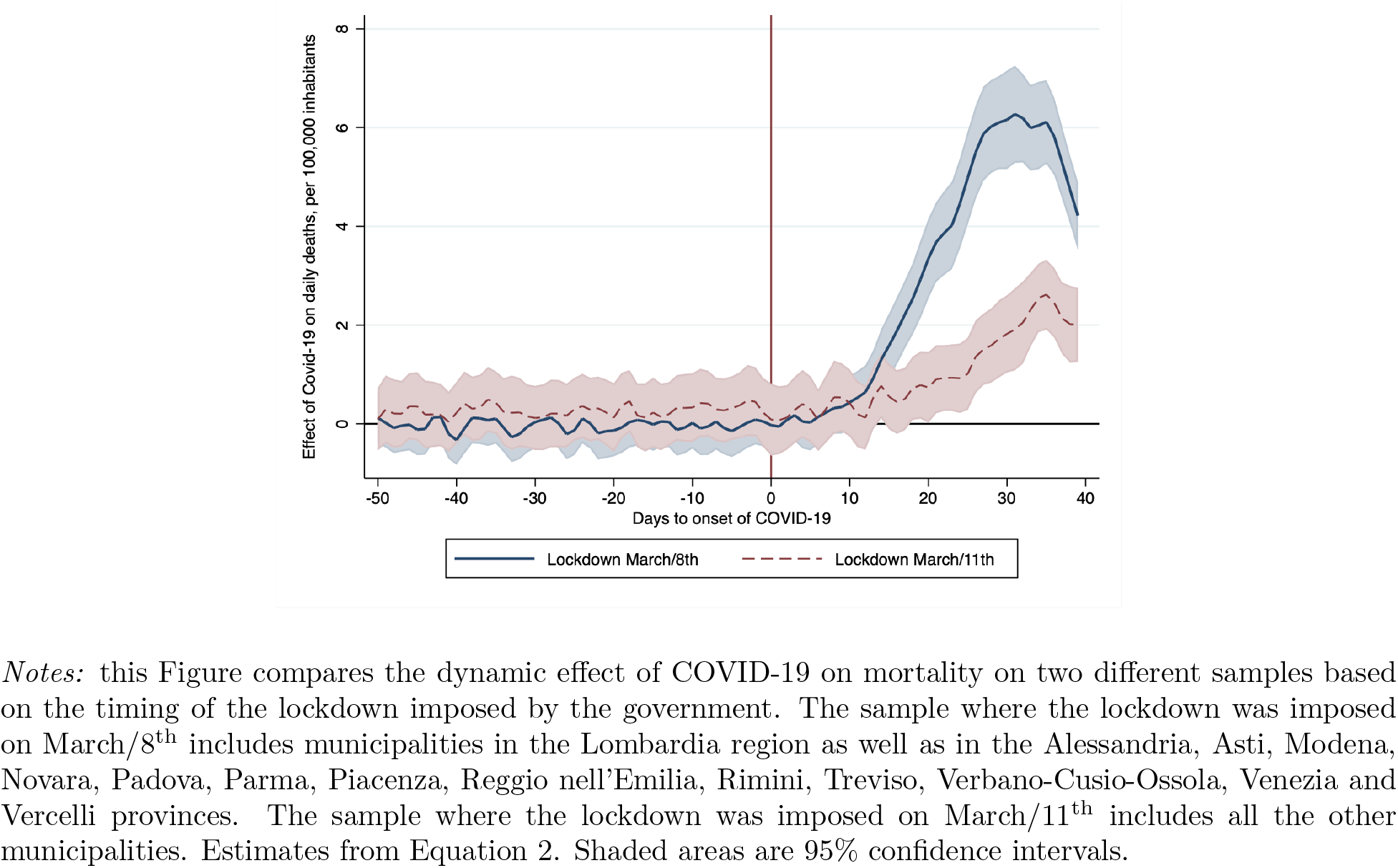
Variation in lockdown timing and peak in mortality

### D Additional Results on Employment Suspension

**Table D1:** Robustness of suspended firms, municipality characteristics and COVID-19 mortality

|  | Full sample |  |  | Within epicenter |  |  |
| --- | --- | --- | --- | --- | --- | --- |
|  | (1)<br>Baseline | (2)<br>Controls | (3)<br>Time sample | (4)<br>Baseline | (5)<br>Controls | (6)<br>Time sample |
| Average effect | 6.00 | 6.10 | 5.56 | 2.81 | 2.83 | 2.32 |
|  | 0.28 | 0.32 | 0.26 | 0.13 | 0.14 | 0.11 |
| Suspended services | −0.80*** | −0.73*** | −0.47** | −0.42*** | −0.39*** | −0.38*** |
|  | 0.25 | 0.24 | 0.23 | 0.11 | 0.11 | 0.10 |
| Suspended factories | −0.06 | −0.05 | −0.12 | −0.01 | −0.02 | −0.07* |
|  | 0.08 | 0.08 | 0.08 | 0.06 | 0.06 | 0.04 |
| Employment rate | 0.50** | 0.61*** | 0.36* | 0.02 | 0.04 | 0.01 |
|  | (0.20) | (0.20) | (0.18) | (0.04) | (0.05) | (0.05) |
| Digital labor | −1.06*** | −1.24*** | −0.75** | −0.22** | −0.23** | −0.14 |
|  | (0.35) | (0.36) | (0.31) | (0.11) | (0.10) | (0.13) |
| External commuting | 1.10*** | 0.76*** | 0.84*** | 0.36*** | 0.43*** | 0.29*** |
|  | (0.24) | (0.27) | (0.21) | (0.10) | (0.11) | (0.11) |
| Population density | 0.17 | 0.06 | 0.13 | 0.72*** | 0.79*** | 0.66*** |
|  | (0.28) | (0.30) | (0.26) | (0.11) | (0.12) | (0.11) |
| Share 80+ | 2.31*** | 1.78*** | 2.19*** | 0.26** | 0.45*** | 0.34*** |
|  | (0.41) | (0.49) | (0.40) | (0.11) | (0.14) | (0.12) |
| High school grads | −1.85*** | −2.30*** | −1.61*** | −1.17*** | −1.08*** | −1.06*** |
|  | (0.44) | (0.40) | (0.40) | (0.14) | (0.13) | (0.14) |
| Income inequality | −0.23 | 0.46 | −0.03 | 0.11 | −0.16 | 0.21* |
|  | (0.41) | (0.37) | (0.37) | (0.10) | (0.12) | (0.11) |
| Share health care | 0.06 | −0.13 | 0.09 | 0.19* | 0.24* | 0.15 |
|  | (0.23) | (0.22) | (0.20) | (0.10) | (0.13) | (0.10) |
| PM10 | 0.79** | 1.05*** | 0.31 | 0.37*** | 0.31*** | 0.36*** |
|  | (0.38) | (0.39) | (0.32) | (0.10) | (0.10) | (0.11) |
| Observations | 137,923 | 137,290 | 124,232 | 549,256 | 533,603 | 483,148 |
| R-squared | 0.14 | 0.14 | 0.13 | 0.03 | 0.03 | 0.03 |

*Notes:* This table reports robustness checks on the effects of business shut downs on mortality. Columns 1 and 4 report our baseline results (see Columns 3 and 6 of Table 1). Columns 2 and 5 report results adding five additional controls: the share of Chinese immigrants, the share of women, an index measuring the quality of sanitary services, an internal commuting index and the share of temporary workers. Columns 3 and 6 report estimates obtained over a longer time sample, spanning until April/15<sup>th</sup>. \* $p < 0.05$ , \*\* $p < 0.01$ , \*\*\* $p < 0.001$ .

### E Additional Results on Emergency Care

**Figure E1:**
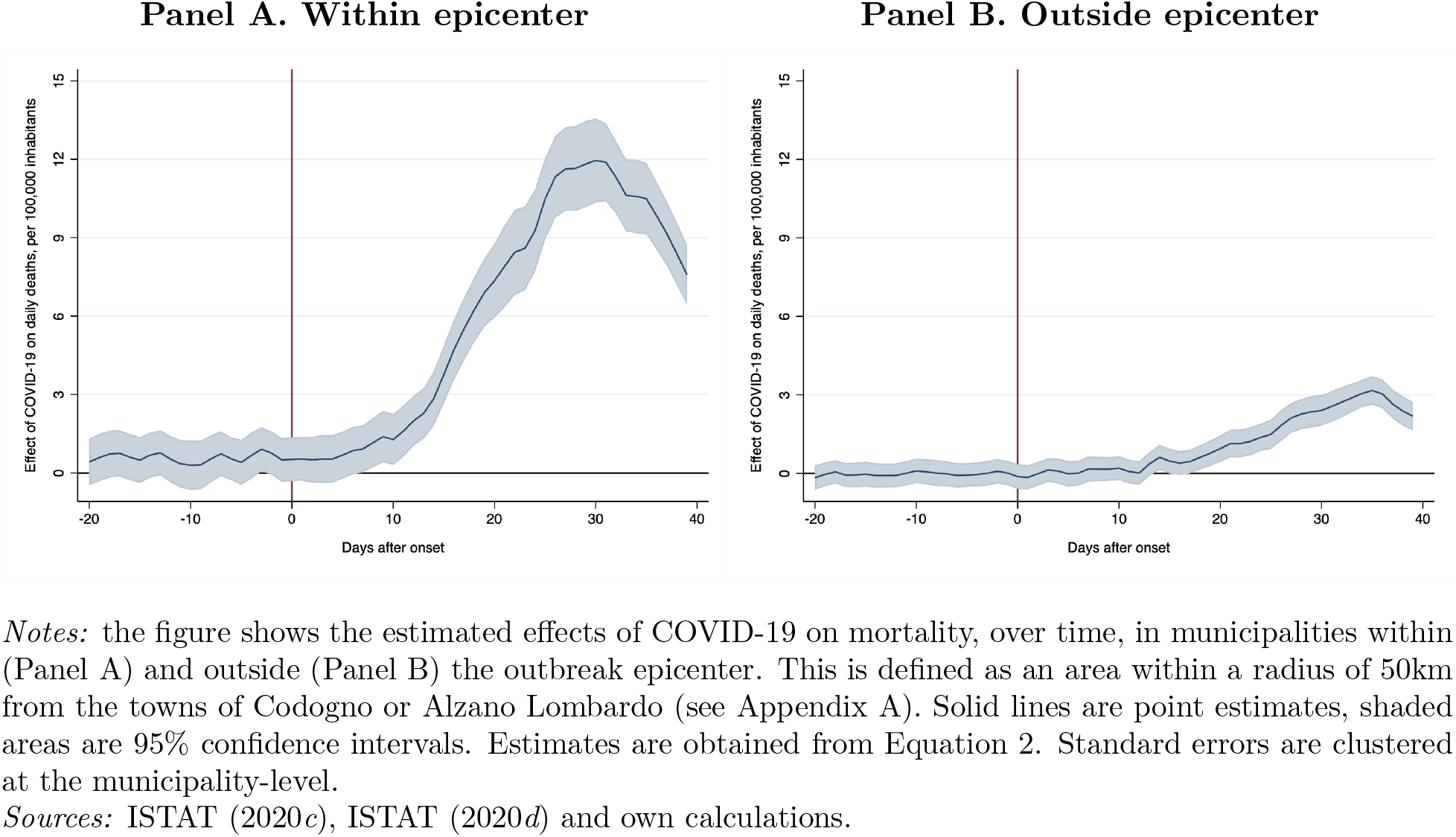
The effect of COVID-19 within and outside the outbreak epicenter

**Figure E2:**
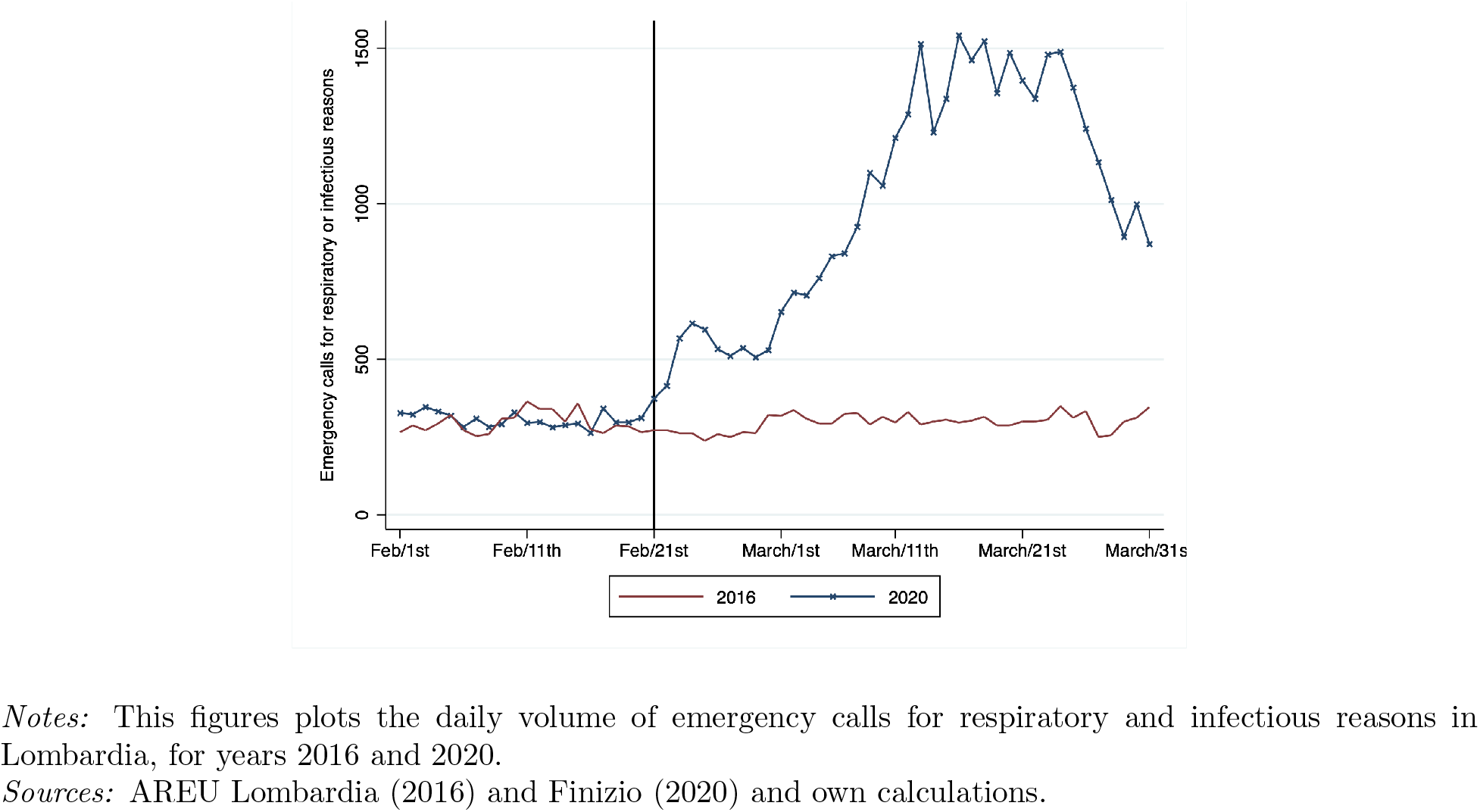
Emergency calls in Lombardia, 2016 and 2020

**Figure E3:**
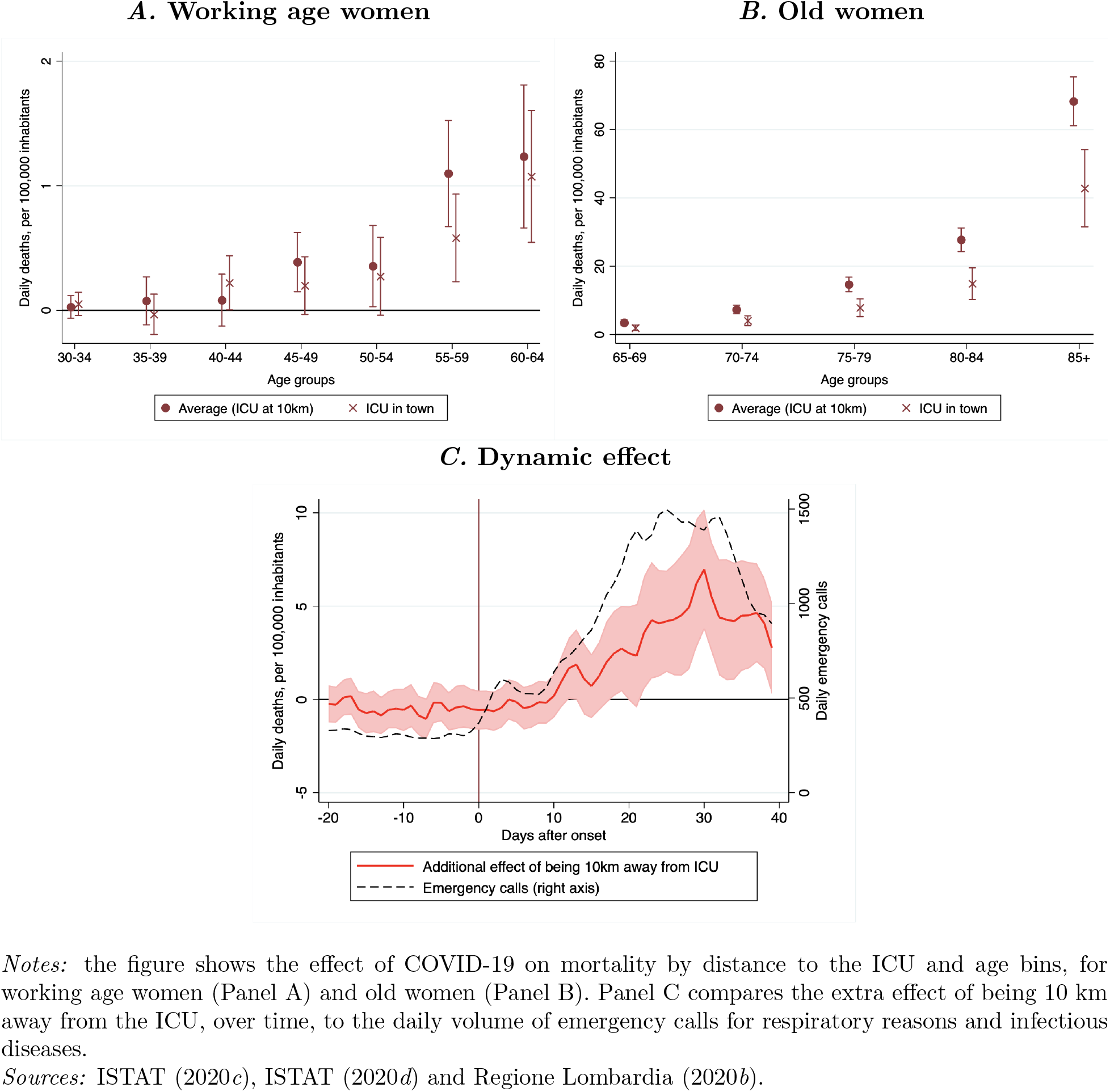
Distance to ICU, overwhelmed system and COVID-19 mortality, women

**Table E1:** Additional effect of COVID-19 on daily deaths per 100,000 people by sex from ICU at 10km

|  | Baseline | At epicenter,<br>controls | Outside epicenter | Outside epicenter,<br>controls |
| --- | --- | --- | --- | --- |
|  | (1) | (2) | (3) | (4) |
| ***Men*** |  |  |  |  |
| ICU at 10km | 2.86***<br>(0.68) | 2.54**<br>(0.77) | 0.55*<br>(0.21) | 0.28<br>(0.29) |
| $R^2$ | 0.07 | 0.07 | 0.02 | 0.02 |
| ***Women*** |  |  |  |  |
| ICU at 10km | 2.61***<br>(0.67) | 2.52***<br>(0.68) | 0.47*<br>(0.22) | 0.37<br>(0.27) |
| $R^2$ | 0.04 | 0.05 | 0.01 | 0.01 |
| Observations | 135,298 | 135,298 | 88,781 | 88,600 |

*Notes:* This table reports the additional effect on mortality from ICU being at 10km from the municipality, by sex. Column (1) reports our baseline specification, i.e., effects at the epicenter with time and fixed effects without additional control variables. Column (2) adds additional controls, by selecting the control variables that are statistically significant in as shown in Table 3: share of suspended services, digital labor, external commuting, population density, share 80+, high school grads, share health care, and PM10. Columns (3) and (4) reproduce this exercise, limiting the sample to municipalities 50km or further from the epicenter. \* $p < 0.05$ , \*\* $p < 0.01$ , \*\*\* $p < 0.001$

### F Additional Results on Nursing Homes

This appendix zooms in on nursing homes and explore whether they may have contributed to the high death toll of COVID-19. This analysis is motivated by the third stylized fact discussed in Section 3 – that the scale of undercounting of COVID-19 deaths is higher among the elderly. Anecdotal evidence suggests that COVID-19 had disproportionately high mortality effects in nursing homes (The Economist, 2020*b*) and, since Italy does not include nursing home deaths in its COVID-19 statistics, there are reasons to believe that they may account for a big chunk of the undetected deaths.

But why are nursing homes particularly exposed to COVID-19? With a large number of residents sharing the same common spaces and having close contacts with multiple staff members, they may have acted as hotbeds of contagion. Moreover, as in Italy nursing homes do not qualify as medical centers, they were heavily understaffed and unprepared to deal with the crisis, lacking protective equipment for staff and emergency care equipment for infected patients (Istituto Superiore di Sanita, 2020). In Lombardia, these inherent characteristics of nursing homes may have been particularly aggravating, as the regional authority decided to relocate COVID-19 positive patients with mild symptoms from hospitals to nursing homes (La Stampa, 2020).

We start by analyzing registry data on nursing homes from Regione Lombardia (2020a) and find that nursing homes are very prevalent in the territory. About 50% of all municipalities have a nursing home in town. Overall, the population of nursing home residents tops 65,000. We then construct a municipality-level variable measuring the proportion of the elderly residing in a nursing home, by age and gender (see Appendix A for more details).^21^ Next, we test whether COVID-19 had an extra effect on mortality in municipalities with a higher share of people living in nursing homes. To do so, we estimate a specification in which we interact the proportion of nursing home residents over the total population with the treatment effect from COVID-19 (similar to what done in Equation 4 with ICU distance). In Table F1 below, we show the mortality effect of COVID-19 of an increase of 10 percentage point in the share of the elderly population living in nursing homes.^22^

Among ages between 70 and 79, we do not find an additional effect on mortality from being in a nursing home for men. For men aged 80 and over, however, the results suggest that living in a nursing home may have significantly increased the chance of dying during the COVID-19 epidemic – by 20 to 30 more daily deaths per 100,000 people (equivalent to about a 30% higher mortality rate). For women, we find additional mortality effects from living in nursing homes already at 70 years old. The effects increase with age, reaching 20 more daily deaths per 100,000 people for those aged 85+ (50% more mortality). Our results are robust to the inclusion of a comprehensive set of control variables (see notes in Table F1).

People over 80 represent two-thirds of the total excess deaths, and our analysis shows that many of them could have been prevented through better preparedness of nursing homes. Providing adequate protective equipment is key to protecting its residents and staff. Even more essential is the need to identify and isolate positive cases, and prevent staff from going to work if they are affected. Ramping up testing capacity among nursing homes residents and staff is thus key to preventing the unfortunate outcomes observed.

**Table F1:** Additional effect of COVID-19 if 10% of 85+ were in nursing homes

|  | Aged 70-74 |  | Aged 75-79 |  | Aged 80-84 |  | Aged 85+ |  |
| --- | --- | --- | --- | --- | --- | --- | --- | --- |
|  | Baseline<br>(1) | Controls<br>(2) | Baseline<br>(3) | Controls<br>(4) | Baseline<br>(5) | Controls<br>(6) | Baseline<br>(7) | Controls<br>(8) |
| **Men** |  |  |  |  |  |  |  |  |
| 10% in N.H. | 2.92<br>(7.61) | 6.27<br>(8.24) | 11.21<br>(8.07) | 13.09<br>(8.94) | 29.31***<br>(7.09) | 29.33***<br>(7.40) | 21.35***<br>(3.63) | 19.72***<br>(3.72) |
| $R^2$ | 0.01 | 0.01 | 0.02 | 0.02 | 0.01 | 0.02 | 0.01 | 0.01 |
| **Women** |  |  |  |  |  |  |  |  |
| 10% in N.H. | 4.90*<br>(2.04) | 4.26*<br>(2.14) | 12.89***<br>(3.06) | 13.45***<br>(3.05) | 13.42***<br>(3.08) | 14.58***<br>(3.22) | 20.89***<br>(3.66) | 19.89***<br>(3.68) |
| $R^2$ | 0.00 | 0.00 | 0.01 | 0.01 | 0.01 | 0.01 | 0.02 | 0.02 |
| Observations | 136,045 | 136,045 | 136,045 | 136,045 | 136,045 | 136,045 | 136,045 | 136,045 |

*Notes:* This table reports the additional effect on mortality when the proportion of people in nursing homes increases in 10 percentage points. Columns (1), (3), (5), and (7) report the baseline specification, i.e., effects at the epicenter with time and fixed effects without additional control variables. Columns (2), (4), (6) and (8) add additional controls, by selecting the control variables that are statistically significant in as shown in Table 3: share of suspended services, digital labor, external commuting, population density, share 80+, high school grads, share health care, and PM10. \* $p < 0.05$ , \*\* $p < 0.01$ , \*\*\* $p < 0.001$

## Footnotes

1 For about 70% of municipalities, the data go up to April/15th. We restrict the main analysis to the January/1^st^-March/31^st^ period and use the rest of the sample for some extensions.

2 To calculate deaths attributable to COVID-19, we compare deaths in 2020 to deaths in 2016 and assume that any excess deaths in 2020 are induced by COVID-19 (see Section 4 for more details). Comparing deaths in 2020 to the average of the five preceding years would lead to a very similar estimate.

3 Our definition of deaths attributable to COVID-19 includes both direct deaths (people dying of COVID-19) and indirect deaths (people dying for causes related to COVID-19, such as overcrowded hospitals, see The Economist, 2020*a*). Indirect deaths are net of fewer deaths resulting from government social distancing policies (for instance, due to fewer road accidents and accidents on the workplace). Through a simple back-of-the-envelope calculation we estimate that indirect deaths might account for 3% to 5% of all deaths attributable to COVID-19 (see Appendix B for more details).

4 In Figure C2 in Appendix C, we show that using the average of the five preceding years as control group leads to very similar estimates.

5 We use population weights to account for the many small municipalities in the sample. Additionally, while in Section 2 we presented graphical evidence for all the municipalities in the sample, for the empirical analysis, we windsorize municipalities with less than 1,000 inhabitants. These account for about 20% of the sample of municipalities but make up for just over 1% of the population. The results would be virtually unchanged if we included all municipalities.

6 To further check the efficacy of the lockdown, we also exploit a small regional variation in the timing of the lockdown, which was first imposed in Lombardia and fourteen other provinces on March/8^th^, and then extended to the entire country, on March/11^st^. We estimate the dynamic specification of Equation 2 on the two samples of municipalities that entered in lockdown on March/8^th^ and March/11^th^ respectively and find that the effect of COVID-19 on mortality peaked a few days earlier in the sample where the lock-down was imposed earlier (Figure C3 in Appendix C).

7 The government policy involved both manufacturing and construction industries. We refer to them as factories for simplicity.

8 The coefficients for the share of employment in overall suspended firms, suspended services, and suspended factories are normalized to take into account the different dummies through which they are estimated. They show the effect of a 10 percentage points increase in the share of employment in suspended firms, on average during the entire epidemic period (not over the post-March/11^th^ or post-March/22^nd^ periods).

9 It may be that we do not find effects of shutting down factories on COVID-19 mortality because the effects take time to materialize and our sample ends on March/31^th^ period. To test whether this is the case, we estimate Equation 3 on the sample up to April/15^th^. This includes less municipalities, but it is still fairly representative (see Footnote 2). The results are in line with the baseline estimates.

10 The results are also robust to clustering the standard errors at the local labor market level (estimates available upon request).

11 Indeed, particularly in Italy’s north, a large chunk of employment in manufacturing and construction industries is concentrated in micro firms, with less than 10 workers per unit. Contact is thus fairly limited and, if infected workers are readily identified, contagion may be effectively contained without having to close them down.

12 When considering only the sample of municipalities within the outbreak epicenter, population density is not significant. This may be due to its relatively low variation in this restricted sample.

13 ICU capacity has been strengthened during the COVID-19 epidemic and these additional emergency ICU beds are not recorded in our data on health facilities, which are as of end-2019. However, this does not impact our distance-to-ICU measure because the expansion of ICU capacity was concentrated in municipalities that already had ICUs in town (mostly in Bergamo, Crema, and Milano).

14 Our distance-to-ICU variable measures distance as a straight line. Accounting for roads and traffic, a trip of 10km as the crow flies may take several tens of minutes.

15 The closest ICU is at 9.2 km in the average municipality. We consider 10 km for simplicity. The effect in municipalities with an ICU in town is simply given by 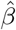, while the effect in those with the closest ICU being at 10 km is given by 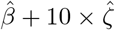.

16 Unfortunately, data on health conditions at the municipality-level is not available.

17 To construct the index we used data on the number of employees in each 2-digit industry from ISTAT (2020a) and the classification of digitalization of work by industry of Manyika et al. (2015). We assign weight 5 to the ICT, media, professional services, and finance and insurance industries, weight 4 to the wholesale trade, utilities, and oil and gas industries, weight 3 to the personal and local services, education and manufacturing industries, weight 2 to the real estate, transportation and warehousing, health care and construction industries, and weight 1 to the retail trade, hospitality, mining and entertainment, and recreation industries. For each municipality, we then compute the weighted sum of employment (using the weights described above) and divide it by overall employment times five. The resulting index assigns similar weights per industry than the index constructed by Dingel and Neiman (2020) using the Occupational Information Network (O*NET) surveys.

18 We acknowledge that ICU capacity has been strengthened during the emergency. These emergency ICU beds are not recorded in the data on health facilities collected. Yet, it should not impact our distance measure, as the expansion in ICU beds happened in municipalities where ICUs were already in place.

19 Almost every nursing home in Lombardy has hundreds of people in their waiting lists. We have calculated that there are, on average, 157 people waiting for every bed that is still available (ATS Insumbria, 2020).

20 PM10 is considered to be above the limit if its day average is above 50 *mg*/*m*^3^ (ARPAE Emilia-Romagna).

21 To construct this variable, we assumed full occupancy. Indeed, almost every nursing home in Lombardy has hundreds of people in their waiting lists. On average, there may be more than 150 people waiting for every bed that is still available (ATS Insumbria, 2020).

22 The average share of 85+ residing in nursing homes (9.4%). For younger ages, the average share living in nursing homes is lower (1.7% for ages 70-74, 2% for ages 75-79, and 4% for ages 80-84).

